# Transcriptomic profiling uncovers mis-splicing and fusion transcripts in ALS

**DOI:** 10.64898/2026.02.05.26345503

**Authors:** Huilin Xu, Tiziana Petrozziello, Adel Boudi, Shota Shibata, Sommer S. Huntress, Eric J. Granucci, Alireza Shahryari, Xuefang Zhao, Maheswaran Kesavan, Ayleen L. Castillo Torres, Ranee Zara B. Monsanto, John Lemanski, Bimal Jana, Celin E.F. De Esch, Merit E. Cudkowicz, James D. Berry, NYGC ALS Consortium, Harrison Brand, Michael E. Talkowski, Ricardo Mouro Pinto, Dadi Gao, Ghazaleh Sadri-Vakili

## Abstract

Advances in transcriptomics have transformed our understanding of amyotrophic lateral sclerosis (ALS), a progressive neurodegenerative disease, revealing disrupted gene expression profiles and highlighting the multi-system biology of ALS. Despite major advances, transcriptomic studies have only begun to capture the complexity and the molecular hierarchy of transcriptomic alterations in ALS. To resolve and characterize the transcriptome in ALS, we performed a comprehensive reanalysis of bulk RNA sequencing from the New York Genome Center ALS Consortium cohort across five post-mortem tissues including motor and frontal cortex, cervical and lumbar spinal cord, and cerebellum. By deploying dual analytical pipelines - one reference-based to model canonical events and one *de novo* to detect transcript structural novelties - we disentangled the quantitative and qualitative architectures of ALS.

Our reference-based analysis revealed that the ALS transcriptome is defined primarily by splicing failure rather than changes in gene expression. Aberrant splicing events, particularly intron retention, outnumbered differentially expressed genes by an order of magnitude. This widespread loss of fidelity disproportionately affected RNA-binding proteins (RBPs), suggesting a collapse in their autoregulatory feedback loops. Deconvolution of these signals identified distinct cellular vulnerabilities: transcriptional disruptions were enriched in glial cells in sporadic cases but in neuronal cells in C9ORF72-positive cases. Furthermore, we observed sex-specific dysregulation, with male individuals exhibiting greater disruption in guanosine triphosphatase signaling and ciliary organization pathways.

In parallel, our *de novo* analysis uncovered a significant burden of ALS-specific RNA fusions that were absent in controls. Whole-genome sequencing of the same individuals, together with a larger reference population, confirmed that disease-specific fusions do not arise from genomic structural variants, indicating a transcriptional rather than genomic origin. PacBio long-read Iso-Seq in an independent post-mortem cerebellar cohort confirmed multiple ALS-specific fusions identified by short-read sequencing. Similarly, a junction-specific BaseScope assay detected one of the fusion transcripts, *FAM69A-EVI5*, in human ALS cerebellum, providing orthogonal evidence for the endogenous presence of an ALS-associated fusion transcript. ALS-specific fusion junctions also displayed a dramatic depletion of RBP motifs relative to canonical splice junctions, and the presence of these sparse disease-specific fusions was strongly correlated with severe splicing outliers in genes governing guanosine triphosphatase activity.

Altogether, our results delineated a transcriptome characterized by aberrant splicing with tissue- and sex-specific changes and identified structural-variant-independent RNA fusions as an additional feature of RNA-processing dysregulation in ALS. This integrated view provides a mechanistic scaffold for the development of splicing-centered and RNA-structural therapeutic strategies for ALS.

**Graphical abstract:** 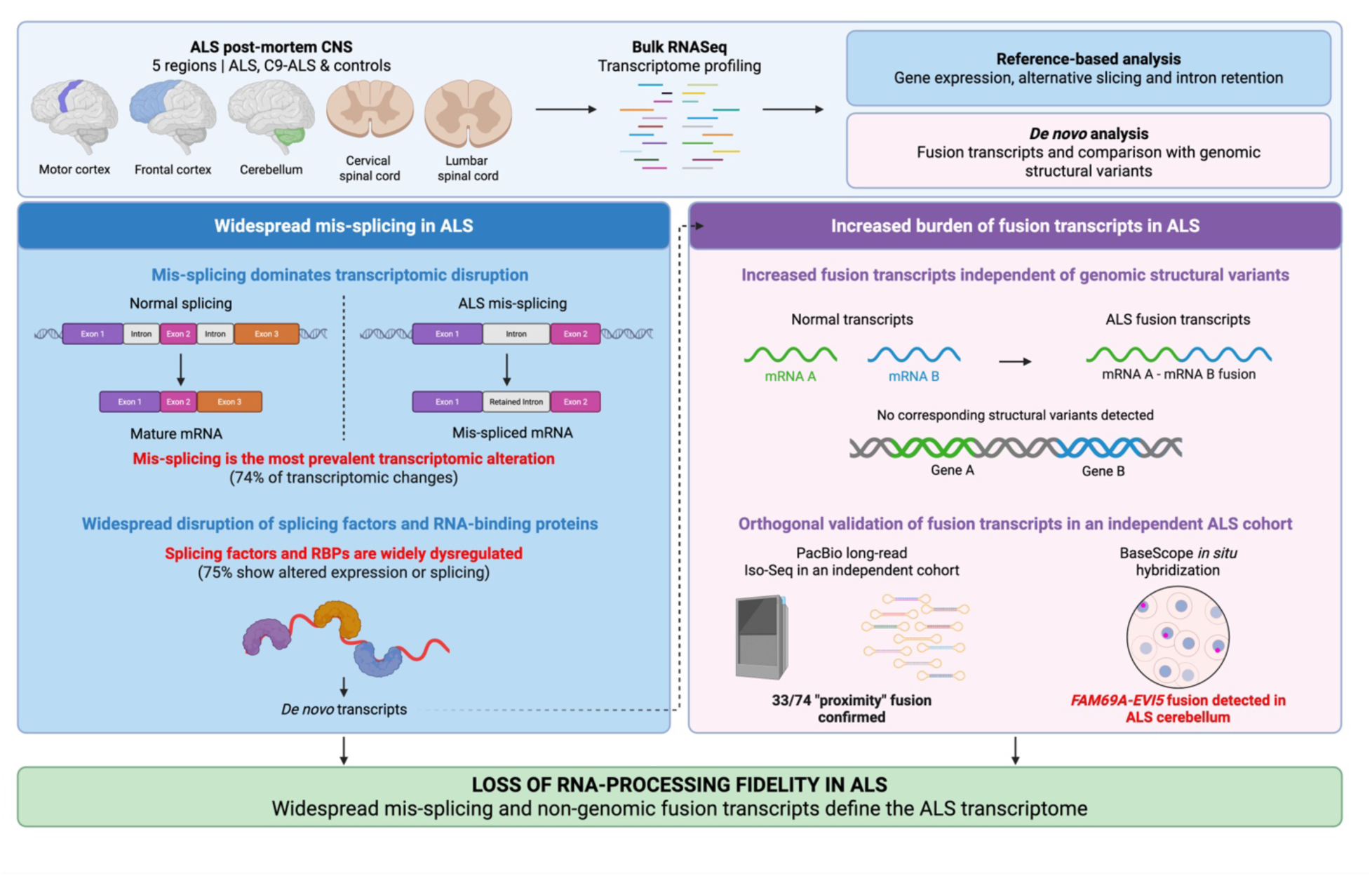

## Introduction

Amyotrophic lateral sclerosis (ALS) is a fatal neurodegenerative disease primarily characterized by the progressive degeneration of motor neurons, leading to muscle weakness, paralysis and respiratory failure.^1,2^ ALS can present as sporadic (sALS) in approximately 85-90% of cases and familial (fALS) in about 10-15% of cases.^3^ Although a variety of genes have been linked to fALS, including superoxide dismutase 1 (*SOD1*), TAR DNA binding protein (*TARDBP*), fused in sarcoma (*FUS*), and a hexanucleotide expansion (G_4_C_2_) in *C9ORF72*,^4,5^ genetic risk factors also contribute to sALS. For example, the G_4_C_2_ repeat expansion in *C9ORF72*, which is the predominant genetic cause of ALS, accounts for about 40% of fALS as well as 5-10% of sALS.^6^ sALS and fALS share other underlying pathogenic mechanisms,^7^ such as TDP-43 proteinopathy, dysregulation of RNA processing and splicing,^8–10^ and aggregation of misfolded proteins, particularly RNA-binding proteins (RBPs).^11,12^

Recent large-scale RNA sequencing (RNASeq) studies in ALS post-mortem central nervous system (CNS) tissues and in patient-derived induced pluripotent stem cell (iPSC) models have revealed widespread transcriptional dysregulation^13–21^ providing new insights into disease mechanisms. Transcriptomic analysis from ALS post-mortem brain and spinal cord tissues from the New York Genome Center (NYGC) ALS Consortium identified three subtypes of ALS characterized by gene expression (GE) signatures related to oxidative stress, glial activation and TDP-43 pathology^13,14,17^ Similar alterations were also reported in ALS post-mortem brain samples from different biorepositories.^19,20^ Finally, RNASeq analysis of ALS iPSC lines has identified alterations in RNA splicing, including enrichment in skipped exon and intron retention (IR).^16^ Together these studies demonstrate that transcriptional dysregulation is a common feature in ALS affecting both GE and RNA splicing across CNS tissues and cellular models.

Although it is now clear that ALS exhibits broad transcriptomic disruptions across the CNS, the relative contribution of GE versus splicing changes and the extent of tissue and sex specificity remain to be fully resolved. It is also unclear to what extent these alterations are shared between ALS and its most common genetic form, C9ORF72-ALS (C9-ALS), although recent work is beginning to elucidate these differences.^20^ Furthermore, whether mis-splicing in ALS results in unannotated transcripts and, if so, whether these represent reproducible disease-associated RNA events that contribute to pathology remains unanswered. Here, we sought to address these gaps by deploying dual analytical pipelines, one reference-based to model canonical events and one *de novo* to detect transcript structural novelties, across five post-mortem regions (motor and frontal cortex, cervical and lumbar spinal cord, and cerebellum) of ALS and C9-ALS samples from the NYGC cohort. Our findings reveal an ALS transcriptome characterized by aberrant splicing with tissue- and sex-specific changes and highlight an increased burden of RNA fusions, supporting previous work demonstrating an enrichment of fusion events in ALS.^21,22^ We further provide independent long-read and orthogonal *in situ* validation of selected fusion events, supporting their biological presence in ALS tissue.

## Materials and Methods

### Sample selection

All RNASeq data analyzed in this study were generated by the NYGC ALS Consortium and provided to our group under a collaborative research agreement. The cohort included 1,986 samples from 557 individuals, including ALS (*n*=443) and controls (*n*=114). CNS tissues with more than 200 available samples were used, including motor (mCTX) and frontal cortex (FC), cervical and lumbar spinal cord, and cerebellum. Only samples with an RNA integrity number (RIN)>5 were included, resulting in 367 individuals (*n*=279 ALS, *n*=88 controls). ALS samples from individuals with unknown *C9ORF72* repeats numbers were excluded. ALS samples harboring a *C9ORF72* repeat expansion ≥30 are referred to as C9-ALS, whereas all other ALS samples are referred to as ALS. Additional sample information is provided in Supplementary Table 1.

### RNASeq preprocessing and fusion detection

Illumina universal adapters were first trimmed, followed by alignment against the human reference genome Gencode v26, also used by genotype-tissue expression (GTEx) release v8, via STAR-Fusion (v1.15.1).^23^ Default STAR-Fusion alignment was used to call fusions, except that only unique mapping with ≤5% mismatches was allowed to increase stringency. STAR (v2.7.11a),^24^ the aligner component of STAR-Fusion, also generated the splice-junction (SJ) counts. Gene and intron counts were generated using HTSeq-count (v0.13.5)^25^ and IRFinder (v1.3.1),^26^ respectively. Alternative splicing (AS) and IR were quantified as percent-of-spliced-in (PSI) from SJ and intron counts, respectively (Supplementary Fig. 1a).

### Modeling Gene Expression (GE)

Genes with median counts-per-million <0.5, were excluded. Sample-wise size factors (SF) were estimated using the DESeq2 (v1.44.0) median-ratio method.^27^ Surrogate variables (SurVar)^28^ were estimated from residuals of log_2_-transformed, SF-normalized gene counts, accounted for diagnosis, sex, disease-by-sex interactions, age at death, and RIN. A generalized linear model (GLM) based on the negative binomial distribution then modeled gene counts as a function of biological variables and SurVars for each tissue (**Equations 1-2**):

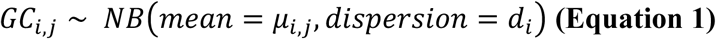

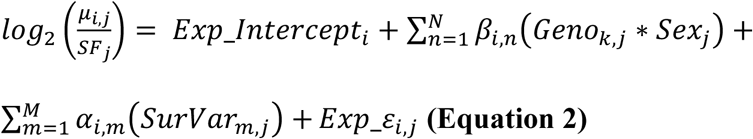

where *Exp_Intercept* represents expression of control female level, *Geno* ALS genotypes, *Sex* male vs female, *β* the weights for biological variables of interest, *α* the weights for SurVars, *ε* the residuals, *i* the *i*-th gene, *j* the *j*-th sample of a tissue, *k* the *k*-th ALS genotype, *n* the *n*-th biological variable of interest, *N* equals 2*k+1*, and *m* the *m*-th SurVar.

### Modeling AS and IR

AS and IR were measured by PSI (Supplementary Fig. 1a) from raw counts at annotated splice junctions in Gencode v26. Samples with <5 reads covering the junction of an event were considered invalid for that event. Events were excluded if <30 valid samples were available in a tissue or <3 samples had PSI values other than 0 or 1. For each tissue, PSI was modeled using Bayesian estimation for a Beta distribution. The probability that PSI was zero or one (i.e. *ρ_i_*) was estimated by intercept-only logistic regression (**Equations 3-5**), followed by estimation of the probability that PSI equaled one (i.e. *λ_i_*) via another intercept-only logistic regression (**Equations 6-8**). PSI values between zero and one were modeled under a Beta distribution, with logit-transformed means represented as a linear combination of biological factors (**Equations 9-10**).

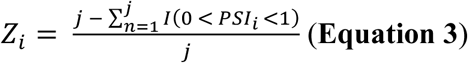

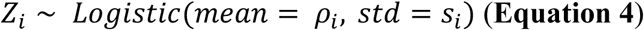

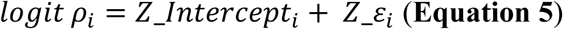

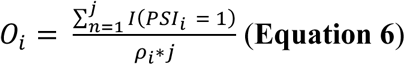

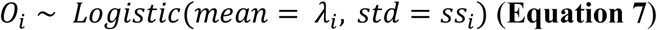

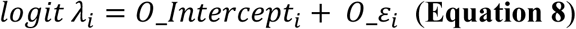

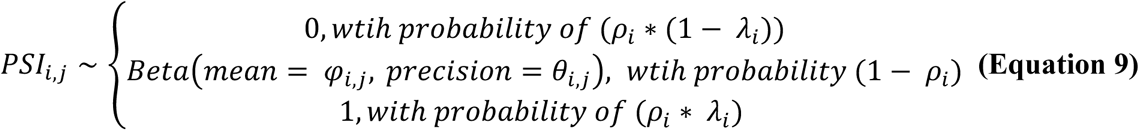

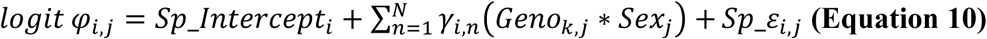

### Functional enrichment test

Gene Ontology (GO) was calculated using Fisher’s exact test against term-wise gene sets and non-redundant host genes of the mis-splicing events, followed by Benjamini-Hochberg false-discovery rate (FDR) correction. Meta-analysis identified GO terms enriched across tissues, followed by semantic similarity clustering.

### Loci comparisons between fusions and SVs

To validate whether STAR-Fusion calls from NYGC RNASeq were potentially derived from structural variants (SVs), we curated candidate SV-derived fusions from whole-genome sequencing (WGS) data from matched NYGC individuals, GTEx, and gnomAD. SVs were called and quality-controlled using GATK-SV as described by Collins et al.^29^ Pipeline and documentation are publicly available online (https://broadinstitute.github.io/gatk-sv/docs/intro). An SV-derived fusion was defined as any SV with breakpoints within two genes such that the rearrangement physically concatenated them. All SV types except breakend (BND) and complex (CPX) were considered. The resulting list was compared with STAR-Fusion calls. Fusions supported by both approaches were considered as SV-derived.

#### Categorizing ALS-specific, control-specific, and shared fusion events

To identify high-confidence ALS-specific fusions, we generated a negative set comprising STAR-Fusion calls from NYGC controls, STAR-Fusion calls from all GTEx brain samples, and all SV-derived fusions except those detected exclusively in ALS NYGC WGS. ALS-derived STAR-Fusion calls overlapping with the negative set were categorized as shared, whereas non-overlapping fusions were considered as ALS-specific fusions. Fusions detected only in NYGC controls were categorized as control-specific (Supplementary Fig. 1b).

### Burden test of fusion events

The number of ALS and control individuals with or without ALS-specific or control-specific fusions, respectively, were counted. Fisher’s exact test assessed whether fusion occurrence was associated with ALS diagnosis in the full NYGC cohort, and in samples with RIN≥5.

### ALS-specific fusion categories

ALS-specific fusions were classified by chromosomal positions and strand orientation (Supplementary Fig. 2) as: (a) inter-chromosomal, where the two parent genes are on different chromosomes; (b) proximity, where the parent genes are on the same chromosome and strand with concordant transcriptional directions (i.e. 5’→3’); (c) discordant orientation (same strand), where the parent genes are on the same chromosome and strand but the fusion is discordant with transcriptional directions (i.e. 3’→5’); and (d) discordant orientation (different strand), where the parent genes are on the same chromosome but opposite strands.

### Association of unexplained transcriptomic variance by ALS genotypes with fusion occurrence

Regression residual models, *Exp_ε_i,j_* (**Equation 2**) and *Sp_ε_i,j_* (**Equation 10**) represented GE and splicing variance not explained by ALS genotypes. GE residuals were tested for association with ALS-specific fusion occurrence using linear regression. For splicing residuals, the association with fusion occurrence was achieved in two steps, by leveraging the uncertainty directly measured from the Bayesian approach. In the first step, the likelihood of zero was calculated from the distribution of response residuals for each sample of each splicing event. A likelihood ≤5% indicated PSI values not fully explained by ALS genotype in that sample and were classified as a splicing outlier. Fisher’s exact test assessed the significance of the odds ratio of having ALS-specific fusion in an outlier sample for each event.

### RBP motif analysis

To generate the query set for RBP analysis, the DNA sequences spanning 100bp flanking each ALS-specific fusion junction identified by STAR-Fusion were extracted. Sequences of 100bp flanking each annotated alternative exon-splice junction formed the background set. RBP motif from the catalog of inferred sequence binding preferences of RBPS (CISBP-RNA) database^30^ were tested for enrichment in query versus background sequences using Simple Enrichment Analysis (SEA) algorithm in the MEME suite.^31^ Motifs with an *E*-value<0.05 were considered significant.

### Independent human post-mortem tissue samples

Post-mortem cerebella were provided by the Massachusetts Alzheimer’s Disease Research Center (ADRC) with approval from the Mass General Brigham (MGB) Institutional Review Board (IRB). Samples were independent of the NYGC cohort. Nine ALS and three control cerebellar samples were assessed. Post-mortem interval (PMI) ranged from 20-47h in controls and 7-54h in ALS. Detailed demographic and clinical information are summarized in Supplementary Table 2.

### RNA extraction and PacBio long-read Iso-Seq

Total RNA was extracted from ALS and control post-mortem cerebellum in TRIzol/chloroform using PhaseMaker tubes following manufacturer’s instruction. Briefly, 100mg of tissue was homogenized in TRIzol (#15596026; Thermo Fisher Scientific, MA), transferred to PhaseMaker tubes, and mixed with chloroform (#2432; Millipore Sigma, MA). Following phase separation, RNA was precipitated in ice-cold isopropanol at -80°C for 30min, washed in 75% ethanol (3x), and resuspended in diethyl pyrocarbonate (DEPC)-treated water. RNA quantity and integrity were assessed using TapeStation (Agilent Technologies, MA), and samples with RIN≥7.0 were advanced for library construction. Poly(A) capture was applied to enrich mRNA, followed by stranded reverse transcription without shearing to preserve full-length cDNA.

For PacBio long-read Iso-Seq, full-length cDNA was generated using the Iso-Seq Express 2.0 kit (Pacific Biosciences; PN: 103-071-500). Twelve barcoded cDNA samples were pooled and converted into a sequencing-ready Kinnex library using the PacBio Kinnex Full-Length RNA kit (Pacific Biosciences; PN: 103-072-000), following the manufacturers’ protocols. Library preparation and sequencing on a PacBio Revio system (Pacific Biosciences, CA) were performed by Broad Clinical Labs (Broad Institute of MIT and Harvard, Cambridge, MA). HiFi reads were processed by Broad Clinical Labs using pbskera, lima, and isoseq refine, including de-concatenation into segmented reads (S-reads), demultiplexing, and extraction of full-length non-concatemer (FLNC) reads with poly(A) trimming. FLNC reads were aligned to the human reference genome GRCh38 (hg38) using minimap2 (v2.28-r1209),^32^ and alignments were sorted and indexed to generate coordinate-sorted BAM files for downstream analyses using samtools (v1.17).^33^

#### Cell culture

Human neuroblastoma SH-SY5Y cells (ATCC® CRL-2266) were cultured in DMEM/F12 medium (Gibco, #11320-033), supplemented with 10% heat-inactivated FBS ((#A5670701, Gibco, ThermoFisher Scientific, MA), 2mM L-glutamine, 50μg/mL streptomycin and 50IU/mL penicillin (#SV30082.01, Cytiva Life Science, MA) and maintained at 37°C in a humidified atmosphere containing 5% CO_2_.

#### SH-SY5Y cells expressing *FAM69A-EVI5*

Lentiviral plasmids encoding the *FAM69A-EVI5* fusion transcript and the corresponding control construct were generated by VectorBuilder: LV[Exp]-EF1A>{FAM69A-EVI5}/FLAG:P2A:mCherry(ns):T2A:Puro (VB241202-1717exf) and pLV[Exp]-EF1A>mCherry(ns):T2A:Puro (VB241202-1740qrn), respectively. Detailed vector maps and sequence information are available through VectorBuilder using the indicated vector IDs. 28,000 cells/well were plated onto 8-well chamber slides (#PEZGS0896, Millipore Sigma, MA), pre-coated with poly-D-lysine hydrobromide (#P6407-5MG, Sigma-Aldrich, MA). Plasmid DNA was introduced using Lipofectamine 2000 (#11668019, ThermoFisher, MA) according to the manufacturer’s instructions and our previously established protocols.^34–36^ Transfections were performed in Opti-MEM (#11058021, Gibco, ThermoFisher, MA) using a final plasmid concentration of 200ng/mL. After 5h, the transfection medium was removed and replaced with fresh complete culture medium. Cells were processed for BaseScope *in situ* hybridization 72h after transfection.

### BaseScope RNA in situ hybridization

RNA *in situ* hybridization was performed using the BaseScope assay (#323970, Bio-Techne, MN) with a custom probe designed to specifically target the junction between *FAM69A* and *EVI5* (#1881901-C1, BaseScope^TM^ probe BA-Hs-FAM69A-EVI5-Junc-C1, Bio-Techne Spatial, MN). The probe consisted of a single ZZ probe pair spanning the fusion junction, thereby restricting signal detection to the *FAM69A-EVI5* fusion transcript rather than either parental transcript. The probe was then used to detect the *FAM69A-EVI5* fusion transcript in transfected SH-SY5Y cells and post-mortem cerebellar tissue from ALS and non-neurological control donors. ALS cerebellum was selected based on prior detection of *FAM69A-EVI5* in the same tissue by PacBio long-read Iso-Seq. BaseScope hybridization and signal amplification were performed according to the manufacturer’s protocol, with sample-specific pretreatment conditions for cultured cells and tissue sections. In brief, cells were fixed in 4% formaldehyde, whereas brain tissues were fixed in 10% neutral-buffered formalin. Following fixation, samples were dehydrated through a graded ethanol series and treated with hydrogen peroxide. For target retrieval and permeabilization, cells were digested with RNAscope Protease III and tissue sections with RNAscope Protease IV. After washing, the custom probe was hybridized to the samples for 2h at 40°C using a HybEZ II oven (Bio-Techne, MN). Following hybridization and washing, sequential signal amplification was performed using AMP reagents 1-8 included in the BaseScope detection kit. Signal was then developed with Fast RED chromogen (1:60 ratio Fast RED-B: Fast RED-A). Samples were counterstained with 50% Gill’s hematoxylin in water and blued with 0.02% aqueous ammonium hydroxide, coverslipped and imaged using an Olympus VS120 slide scanner (Olympus, Evident Scientific, MA). Fusion-specific signal was defined as discrete puncta corresponding to detection of the *FAM69A-EVI5* junction.

### Western blotting

Western blots were performed using previously described protocols.^37–39^ Briefly, 50μg of proteins was separated on a 4-20% Bis-Tris gel and transferred to a PVDF membrane using iBlot Dry Blotting System (Invitrogen, ThermoFisher, MA). Membranes were blocked with 5% milk in TBST and incubated overnight at 4°C with antibodies against ELAVL2 (1:500; #14008-1-AP, Proteintech, IL), and GAPDH (1:1000; #MAB374, Millipore Sigma, Termecula, CA). Membranes were incubated with HRP-conjugated goat anti-rabbit (1:2000; #111-035-003, Jackson ImmunoResearch Laboratories, PA). Proteins were visualized using enhanced chemiluminescence (ThermoFisher Scientific, MA) and quantified using Image J (NIH, Bethesda, MD).

### Statistical analysis

All bioinformatics statistical analyses were performed in R (v4.4.1). To establish DEGs, a Wald test was applied to *Geno* in the GLM (**Equation 2**) to generate a *p*-value for each gene. To establish DAR and DIR, the Bayesian estimation generated the empirical cumulative distribution function (ECDF) of *Geno* (**Equation 10**) for each splicing event. The ECDF at 0 was elevated. If the probability ≤0.5, it was treated like a *p*-value. Otherwise, (1 - probability) was treated as a *p*-value. For the tissues with multiple non-C9 ALS subtypes, a *meta-p* analysis was applied to summarize the significance of those subtypes into an overall ALS group. For DEG, the FDR correction was applied to the *meta-p* values for the overall ALS group and the nominal *p*-values for C9-ALS. A DEG is defined as FDR <0.05. For DAS and DIR, the Bonferroni correction was applied to the *meta-p* values of the overall ALS group and the nominal *p*-values of the C9-ALS group. A DAS or DIR is defined as Bonferroni-corrected *p*-value <0.05.

Functional enrichment analyses were performed using Fisher’s exact test with a significance threshold of FDR<0.1. To identify robust cross-tissue signatures, a *meta-p* analysis was employed with a significance of FDR-adjusted *meta-p* value<0.05. Cell marker enrichment analyses were performed using Fisher’s exact test with a significance threshold of FDR<0.1. The association between splicing outliers and fusion events was evaluated via outlier analysis using Fisher’s exact test, with significance defined as an FDR adjusted *p*-value<0.05. Statistical analyses for protein level assessment were performed using GraphPad Prim. Comparison between two groups were performed using Mann-Whiteny U test; correlations between ELAVL2 levels and disease duration were performed using Spearman correlations. All tests were two-sided, and statistical significance was set at p<0.05. Exact *p* values are reported throughout.

### Study approval

All human samples used in this study were de-identified. The collection, storage and sharing of these samples was approved by the MGB IRB (Boston, MA).

## Results

### Mis-splicing is the primary driver of transcriptomic disruptions in ALS

To define the landscape of transcriptomic dysregulation in ALS, we first quantified the relative contributions of changes in GE and splicing between ALS and control samples across mCTX and FC, cervical and lumbar spinal cord, and cerebellum. Our models revealed that the ALS transcriptome is characterized by a profound loss of splicing fidelity. There were 4,203 (30%) genes harboring both DEGs and mis-splicing (Supplementary Fig. 3a). We identified 1,327 DEGs in mCTX, 3,927 in lumbar spinal cord, 1,612 in cervical spinal cord, 775 in FC and 239 in cerebellum, with the greatest disruption in lumbar spinal cord (Supplementary Table 3, Supplementary Fig. 3b). In contrast, DAS and DIR analyses uncovered 16,921 (in 6,940 genes) mis-spliced events in mCTX, 15,845 (in 6,715 genes) in lumbar spinal cord, 22,142 (in 8,015 genes) in cervical spinal cord, 33,757 (in 9,506 genes) in FC, and 10,425 (in 4,882 genes) in cerebellum out of a total of 608,247 events, with the greatest disruptions in FC (Supplementary Tables 4-5, Supplementary Fig. 3c). Collectively, 54.6% (13,971 genes) of expressed genes exhibited significant transcriptional alterations in at least one CNS region in ALS (Fig. 1a, Supplementary Fig. 3b), while 45.4% (11,618 genes) of genes were unimpacted. Aberrant IR accounted for 48.8% of transcriptomic changes, exceeding both DEG (26%) and DAS (25.2%, Fig. 1b, Supplementary Fig. 3c). Furthermore, 28.4% of disruptions were shared across cortex (motor and frontal) and spinal cord (cervical and lumbar) (Fig. 1c), whereas 2.2%-8.1% were unique to a single region. Such convergent and region-specific patterns might help interpret the heterogeneity of the disease. Hence, we interrogated functional dysregulations across brain regions and according to the presence of the *C9ORF72* expansion (Supplementary Table 1). GO clustering identified six domains of functional deficiency in at least one brain region (Fig. 1d): innate immune response, receptor-mediated immune regulation, phospholipid biosynthesis, neurite development, cytoskeleton organization and protein assembly, and small GTPase signaling regulation (Fig. 1d, Supplementary Table 6). GTPase signaling emerged as a convergent functional signature between ALS and C9-ALS. While C9-ALS demonstrated a severe immune response in the spinal cord, ALS exhibited phospholipid biosynthesis, neurite development and cytoskeleton organization in cortex and spinal cord (Fig. 1d).

**Figure 1.**
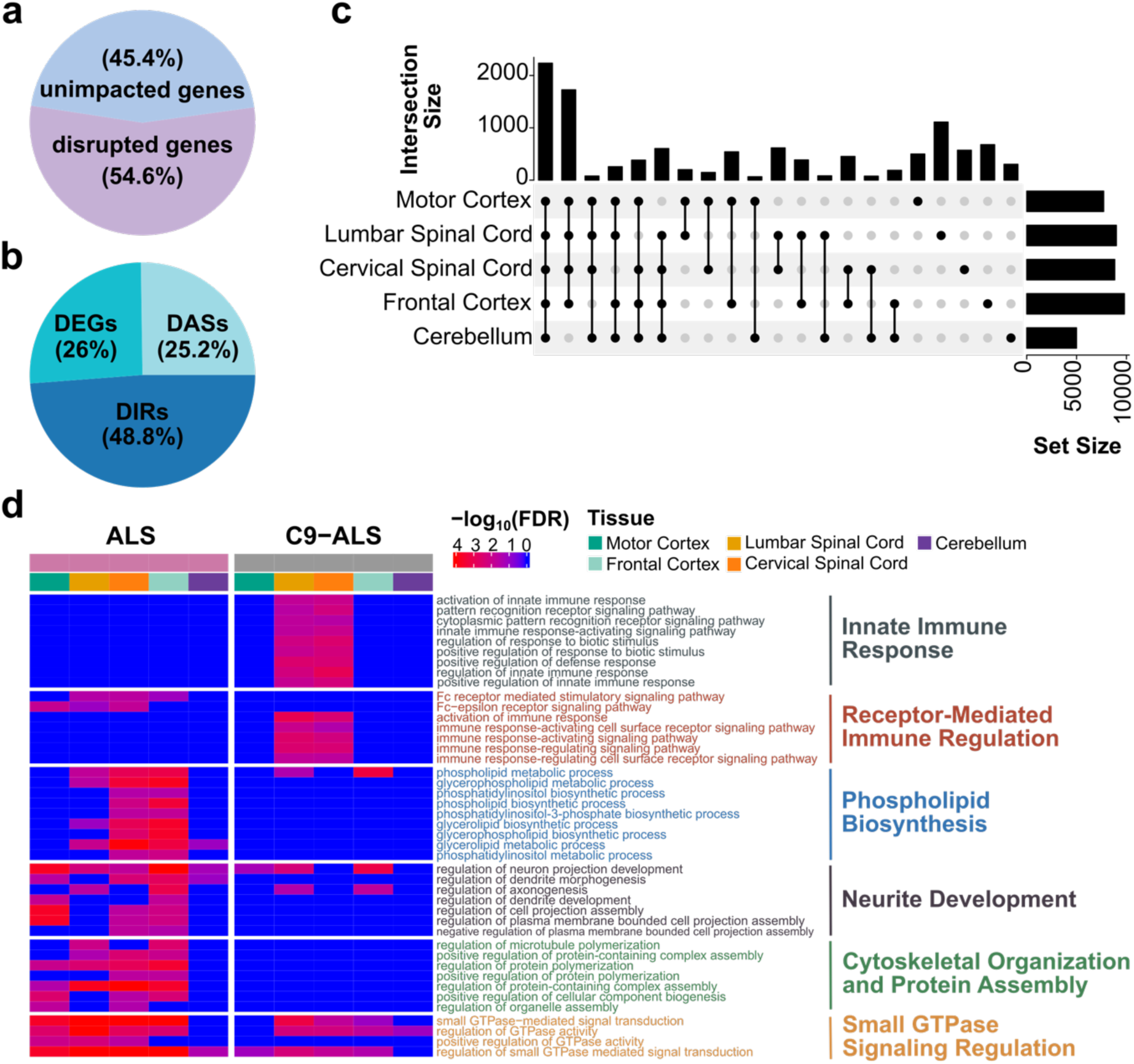
Global transcriptional disruptions in ALS across tissues. **(a)** Proportion of disrupted (54.6%) and unimpacted genes (45.4%) relative to the total number of expressed genes across five tissues, including motor and frontal cortex, cervical and lumbar spinal cord, and cerebellum. **(b)** Proportion of disrupted genes classified as exhibiting DASs (25.2%), DIRs (48.8%), or DEGs (26%). **(c)** UpSet plot depicts the number of genes that are either mis-spliced (i.e., DIR or DAS) or as DEGs across the examined tissue. Horizontal bars represent the total count of affected genes in each tissue, while vertical bars represent the size of each intersection, corresponding to genes shared exclusively between tissue combinations. **(d)** Functional enrichment of disrupted genes, either mis-spliced or DEGs, in each tissue. GO analyses were performed for each tissue across the two ALS subtypes: ALS and C9-ALS. Heatmap color intensity represents enrichment significance (-log_10_FDR), with darker red indicating a larger significant enrichment. GO terms were grouped into clusters based on semantic similarity (*right*) and color-coded accordingly. Displayed clusters contain a minimum of four GO terms, each meeting a *meta-p* value significance (*meta-p* < 0.05) across five tissues and showing significance in at least two tissue types within at least one ALS subtype.

To disentangle contributions of GE and mis-splicing, we performed GO enrichment using both DEGs alone and the combination of DEGs and the host genes harboring DAS/DIR events. We then quantified the impact of mis-splicing by calculating the ΔSignificance, defined as the difference in enrichment significance between the combined set and the DEGs-only set. In cortex, small GTPase signaling, cytoskeleton organization/protein assembly, neurite development, and phospholipid biosynthesis were supported by both DEGs and mis-splicing host genes (Supplementary Fig. 4). In spinal cord, immune pathway enrichment was largely attributable to DEGs and was attenuated by inclusion of mis-splicing host genes (Supplementary Fig. 4). These modality-specific patterns were broadly consistent between ALS and C9-ALS, suggesting shared pathway-level dysregulation across genetic groups.

### Mis-splicing affects splicing factors and specific cell types

Given the predominance of IR in ALS, we next examined molecular factors associated with loss of splicing fidelity. We reasoned that widespread transcriptomic alterations might be self-perpetuating through defects in the RNA splicing machinery. To test this, we interrogated the transcriptional integrity (i.e. either as DEG or mis-spliced) of 396 known splicing factors and RBPs. 297 (75%) of these factors were dysregulated (Fig. 2a, Supplementary Fig. 5), with a significant dependency on ALS. The dominant mode of dysregulation was DAS and DIR rather than DEG, consistent with disruption of autoregulatory feedback loops utilized by many splicing factors, including TDP-43, FUS, and heterogeneous nuclear ribonucleoproteins (hnRNPs). ^40–42^ Most RBP disruptions in ALS exhibited an intensive “inclusion” trend, with increased exon/intron usage (Fig. 2a, Supplementary Fig. 5). *FUS* showed increased IR in all tissues except cerebellum (Fig. 2a), whereas *TARDBP* exhibited increased IR and expression exclusively in lumbar spinal cord (Fig. 2a). C9-ALS instead showed a milder “exclusion” trend (Fig. 2a, Supplementary Fig. 5).

**Figure 2.**
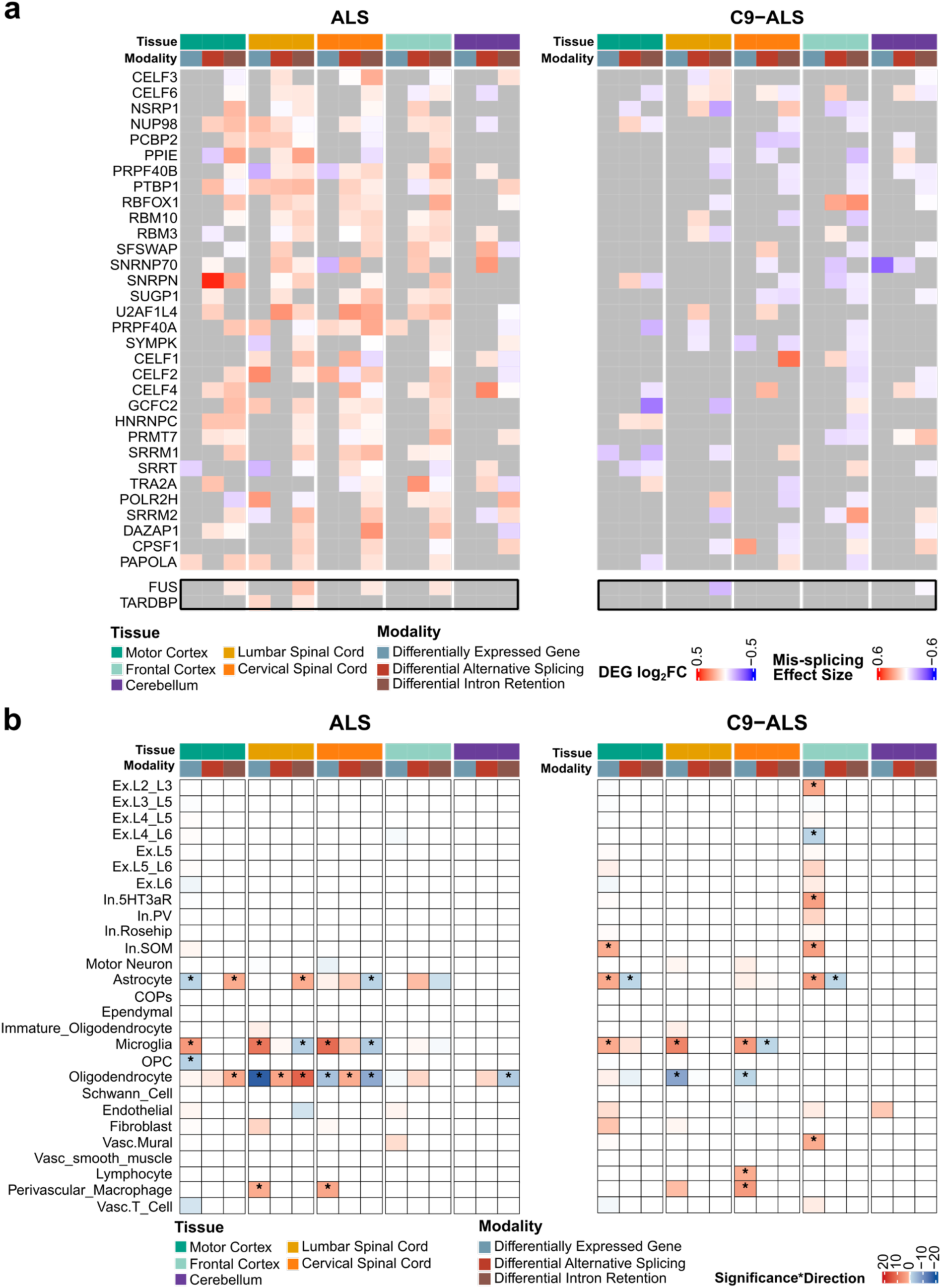
Transcriptional disruptions in ALS impact multiple cell types and are enriched for splicing factors. **(a)** The heatmap displays significant enrichment of transcriptional disruptions for RBPs in ALS (*left*) and C9-ALS (*right*). The top annotation bar includes two groups: the first row indicates tissue type (motor cortex in teal, lumbar spinal cord in yellow, cervical spinal cord in orange, frontal cortex in cyan and cerebellum in purple), while the second row denotes transcriptional modality (DEGs in grey, DASs in red, and DIRs in brown). Color intensity reflects effect size for each ALS subtype compared to controls, with darker red indicating a larger effect size. For DEGs, effect size corresponds to the log_2_(Fold Change) in expression, whereas for DASs and DIRs, effect size corresponds to the maximum absolute ΔPSI value among all mis-spliced events within each gene. **(b)** The heatmap displays enrichment of transcriptional disruptions among cell-type marker genes in ALS (*left*) and C9-ALS (*right*). The top annotation bar comprises two groups: the first row indicates tissue type, while the second row highlights transcriptional modality, as described in panel (a). Red indicates a positive median for log_2_(Fold Change) in expression or ΔPSI value of mis-splicing, whereas blue indicates a negative median for log_2_(Fold Change) or ΔPSI value and white indicates no enrichment for cell type. Color intensity reflects the significance of enrichment, and asterisks denote statistically significant enrichment. * FDR < 0.1.

Given that previous studies suggested cell-type-specific GE changes in ALS, specifically blurring the identity of glial cells in the spinal cord,^14^ we sought to examine to what extent this is associated with mis-splicing. We harmonized cell type annotations across five brain regions and then called cell-type markers using single-cell RNASeq of matched tissues,^43–45^ followed by interrogating their transcriptional integrity. In ALS spinal cord, DEG enrichment confirmed microglial upregulation and oligodendrocytic downregulation, ^14^ with additional enrichment for perivascular macrophages (PVMs) in lumbar and cervical spinal cord (Fig. 2b). In C9-ALS, FC exhibited enrichment for excitatory subtype markers in addition to shared glial alterations found in ALS, thus highlighting a distinct transcriptional signature in C9-ALS. In C9-ALS, PVM and lymphocyte markers were upregulated in cervical spinal cord, while mural markers were upregulated in FC (Fig. 2b). There tended to be no marker enrichment driven by mis-splicing alone in either ALS or C9-ALS (Fig. 2b).

#### Sex-specific transcriptomic disruption in ALS

Several population studies have consistently reported a higher incidence of ALS and shorter survival among men,^46–48^ Building on this evidence, we examined an additive male effect on transcriptomic dysregulations in our models. Male individuals exhibited a greater disruption of GTPase-related signaling and cilium organization in both ALS and C9-ALS (Fig. 3), and a greater disruption in axon and dendrite morphogenesis in ALS. These effects appeared to be tissue-specific: GTPase-related dysregulation was predominantly observed in spinal cord across ALS subtypes, alterations in cilium organization were evident in all regions, and abnormalities in axon and dendritic morphogenesis were confined to ALS spinal cord (Fig. 3).

**Figure 3.**
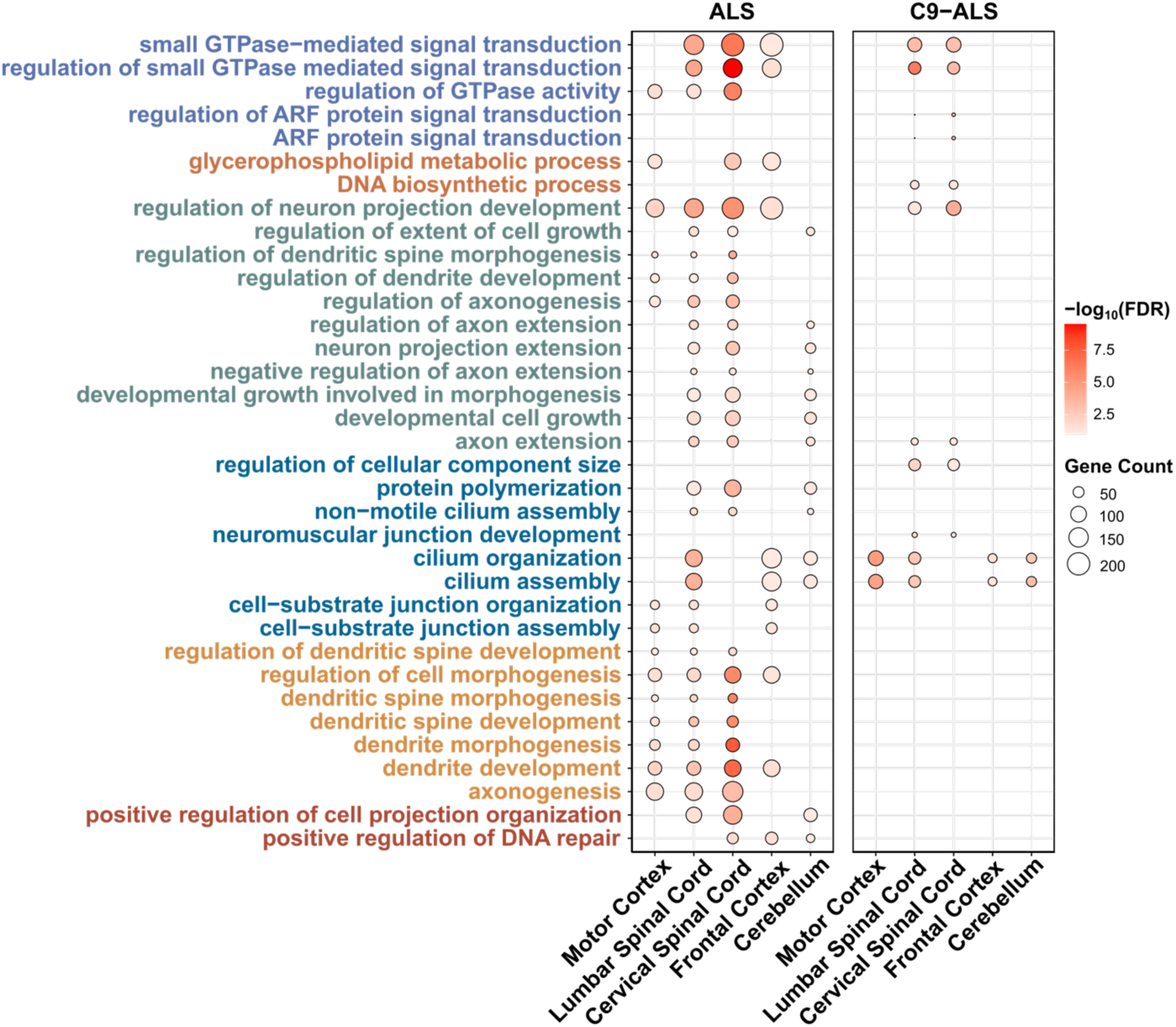
Functional enrichment of male-specific transcriptional disruptions in ALS across tissues. GO enrichment dot plot of male-specific disrupted across tissue types in ALS and C9-ALS. Dot color intensity represents the statistical significance of enrichment measured in -log_10_ (FDR), with more intense colors indicating higher significance, and dot size indicates the number of genes enriched in each GO term. GO terms are grouped and color-coded into clusters based on semantic similarity (*left*).

#### RNA fusions form a significant burden in ALS

Given pervasive aberrant splicing, we examined whether de novo transcriptional events beyond the reference transcriptome were associated with ALS-related dysregulation. Our group previously reported a significant enrichment of gene fusion events in ALS using a subset of the NYGC cohort,^22^ linking gene fusions, a well-established cause of cancer,^49–51^ to neurodegeneration for the first time. With full-cohort and tissue-specific GE and splicing profiles established, we sought to better characterize these fusions and their relationship with ALS-associated transcriptomic changes.

STAR-Fusion calls were categorized as control-specific, ALS-specific or shared. To minimize false positives, we used a negative control set comprising NYGC cohorts and GTEx brain RNAseq fusion together with WGS-derived fusions from NYGC controls, GTEx and gnomAD v4 (Supplementary Fig. 1b). We identified 156 ALS-specific, 51 control-specific, and 359 shared fusion events. Although individual fusion events were rare (Supplementary Table 7), fusion burden was significantly increased in ALS (Fig. 4a) and remained significant in samples with RIN≥5, reducing concern for RNA degradation bias in post-mortem samples (Fig. 4a). However, ALS-specific fusion detection was not strongly associated with tissue (Supplementary Fig. 6a).

**Figure 4.**
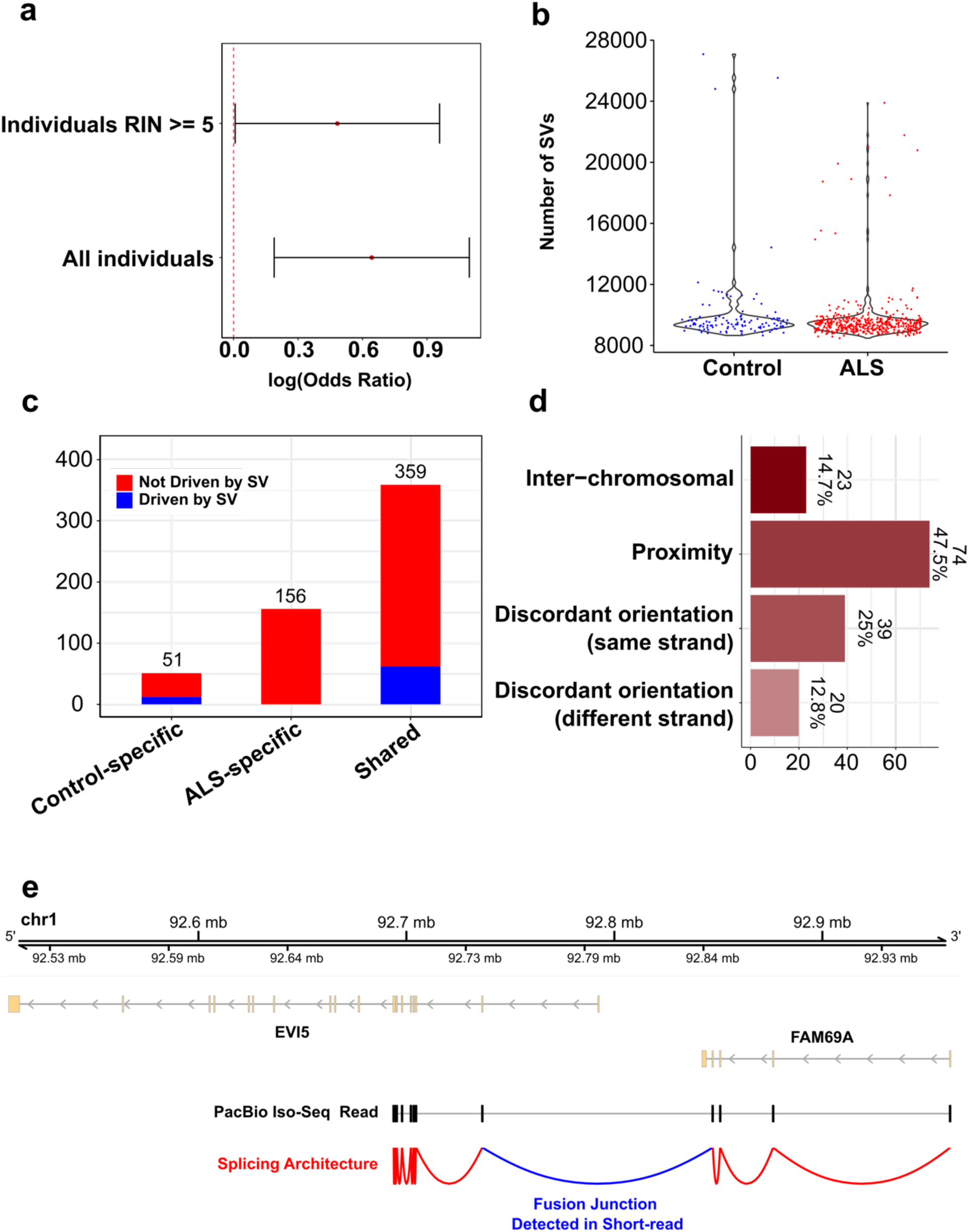
ALS individuals exhibit a significant burden of RNA fusions. **(a)** Error bar plot shows the log (Odds Ratio) of burden test of fusions performed in two ways: using individuals with RNA integrity numbers (RIN) ≥ 5 (*top*; log odd ratio = 0.48) and using all individuals in the NYGC cohort (*bottom*; log odd ratio = 0.64). **(b)** Violin plot depicts the number of SVs observed in control and ALS samples from WGS in the NYGC cohort. **(c)** Stacked bar plot showing the number of control-specific fusions, ALS-specific fusions, and shared fusions attributed to SVs. Fusion events potentially driven by SVs are indicated in blue (control-specific: *n* = 12; ALS-specific: *n* = 0; shared: *n* = 62), whereas fusion events not driven by SVs are shown in red (control-specific: *n* = 39; ALS-specific: *n* = 156; shared: *n* = 297). **(d)** Horizontal bar plot showing the number of ALS-specific fusions in each event according to the chromosomal location and strand orientation of the two fused genes: inter-chromosomal (*n* = 23; 14.7%), proximity (*n* = 74; 47.5%), discordant orientation (same strand) (*n* = 39: 25%), and discordant orientation (different strand) (*n* = 20; 12.8%). **(e)** PacBio Iso-Seq long-read in control and ALS post-mortem cerebellum from the MGH cohort illustrating ALS-specific fusion junctions previously identified by short-read RNASeq in the NYGC cohort. The figure shows the ALS-specific fusion between *FAM69A* and *EVI5* as an example. *FAM69A* and *EVI5* exons are represented in yellow, with arrows (*top*) indicating their transcriptional direction. The PacBio long-read alignment is shown in black exons (*middle*). The sashimi plot depicts the splicing architecture of the long-read alignment, with the blue arch representing the junction detected by short-read RNASeq and the red arches indicating the extended splicing detected in long-read (*bottom*).

Pairing NYGC WGS with RNASeq data allowed us to further investigate whether gene fusions primarily result from DNA rearrangements, such as SVs. On average, SV frequency did not differ between ALS and control genomes (Fig. 4b). None of the 156 ALS-specific fusions was related to known SVs, estimated from >800,000 background genomes (Fig. 4c), whereas at least 18% of shared or control-specific fusions were potentially SV-derived (Fig. 4c). These findings support mechanisms other than DNA rearrangement, such as RNA mis-splicing and/or trans-splicing, as potential contributors to ALS-specific fusion formation.

We further scrutinized the relative orientation and genomic distances between the parent genes of ALS-specific fusions (Supplementary Fig. 2). Among ALS-specific fusion, 23 (14.7%) were inter-chromosomal, 74 (47.5%) were proximity fusions on the same chromosome with their parent genes more than 5kb away, and 59 (37.8%) involved genes on the same chromosome but in discordant orientation, including 39 (25%) on the same strand and 20 (12.8%) on opposite strands, suggesting the potential formation of circular RNA structures (Fig. 4d). While some ALS-specific fusions were only detected in ALS (Supplementary Fig. 6b), we attribute this cautiously to the small sample size of C9-ALS cases. No fusion orientation was strongly associated with ALS subtypes (Supplementary Fig. 6c).

To validate short-read fusion calls, we applied PacBio long-read Iso-Seq to an independent cohort of ALS and control post-mortem cerebellar samples (Supplementary Table 2). Among 74 short-read ALS-specific fusions classified as “proximity” events, 33 (44.6%) fusions were confirmed in ALS samples with the same fusion junction in the long-read alignment and were absent in controls (Fig. 4e, Supplementary Fig. 6d), supporting the reliability of the fusion calls. The ALS-specific fusion between *FAM69A* and *EVI5* is reported as an example in Fig. 4e.

*FAM69A-EVI5* fusion was subsequently used for orthogonal BaseScope validation by designing a junction-specific probe targeting the fusion transcript. Probe specificity was first evaluated in transiently transfected SH-SY5Y cells in which discrete junction-specific puncta were detected in cells expressing *FAM69A-EVI5* but were absent or minimal in cells expressing the mCherry control (Fig. 5a). In post-mortem cerebellum, BaseScope signal was detected in an ALS sample positive for *FAM69A-EVI5* as measured by long-read sequencing, whereas no signal was detected in the control tissue (Fig. 5b), supporting the endogenous presence of the fusion transcripts in human ALS tissue.

**Figure 5.**
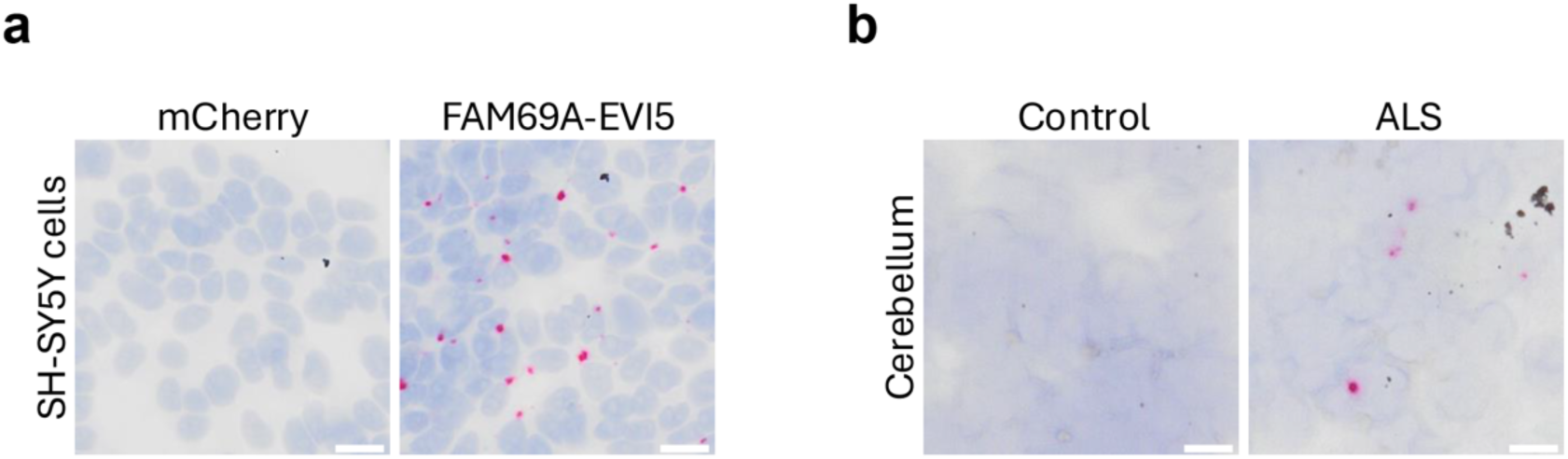
*FAM69A-EVI5* is detectable in human ALS tissue. (a) Representative BaseScope images of SH-SY5Y cells transiently expressing mCherry or *FAM69A-EVI5*. A junction-specific probe was used to detect the fusion transcript; signal are shown in magenta. (b) Representative BaseScope images of post-mortem cerebellum from a non-neurological control and an individual with ALS positive for the *FAM69A-EVI5*. Scale bar 10 μm.

#### Dysregulation of specific RBPs may contribute to ALS-specific fusion

We hypothesized that ALS-specific fusion formation could be related to global mis-splicing, particularly affecting RBP integrity. To assess this, we sought to identify RBPs meeting the following two criteria: specific to fusion events and disrupted in ALS. We analyzed sequences flanking ALS-specific fusion junctions for RBP-binding motifs, providing insight into potential regulatory mechanisms associated with fusions in ALS. Among 98 RBPs with annotated motifs,^52^ 14 were significantly more frequent at fusion junctions compared to canonical splice sites. Twelve RBPs, including *SYNCRIP*, *RBM46*, *PABPN1*, *PABPC4*, *HNRNPR*, *ELAVL2*, and *CPEB4*, also showed transcriptional disruption (either as DEGs or mis-spliced) between ALS and control in at least one CNS region (Fig. 6a, Supplementary Fig. 7a). Among seven fusion-enriched RBPs that were also DEGs, most displayed similar patterns across tissues between ALS and C9-ALS (Fig. 6a). For instance, *ELAVL2* was increased in mCTX from ALS and C9-ALS compared to controls, whereas *CPEB4* levels were decreased in lumbar spinal cord; *SYNCRIP* levels were increased in ALS FC but not in C9-ALS.

**Figure 6.**
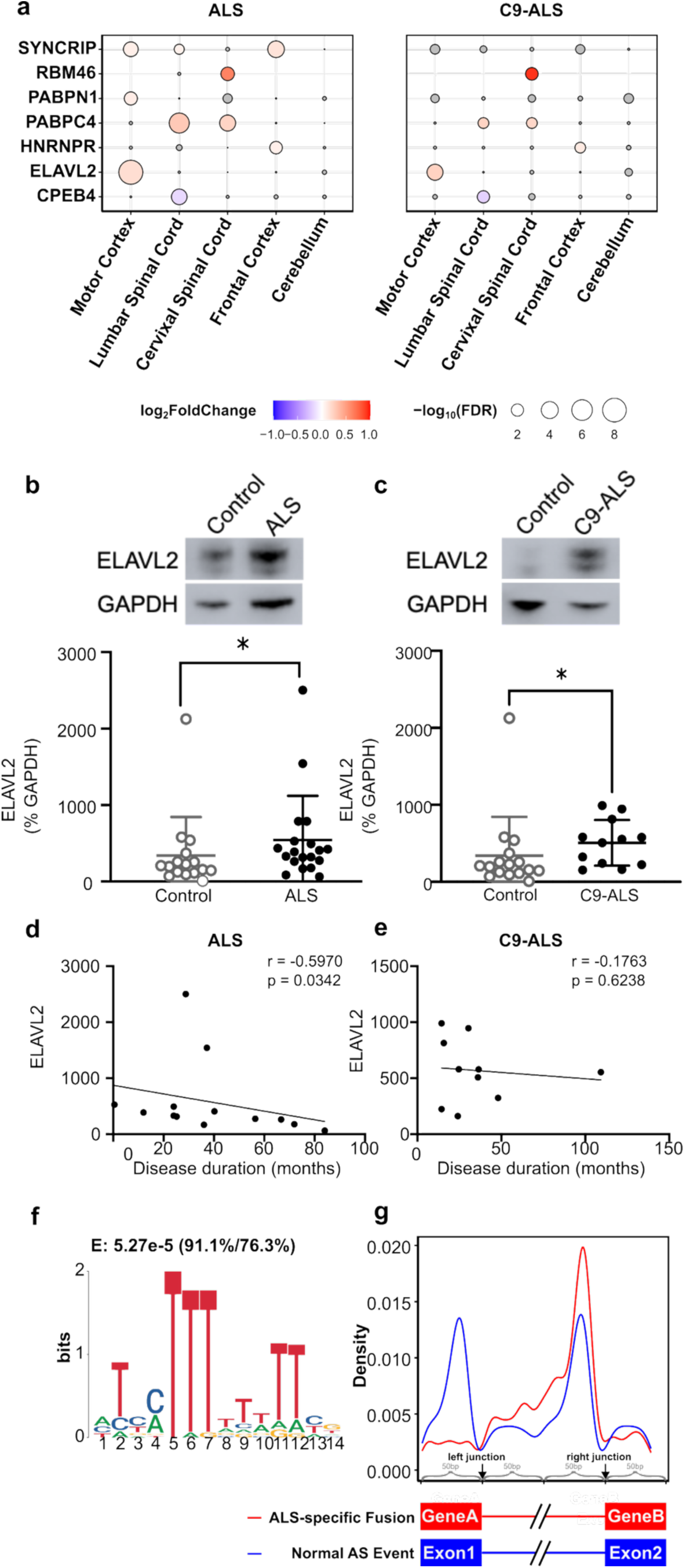
RBP dysregulation potentially contributes to the formation of ALS-specific fusion. **(a)** Dot plot showing significant expression changes of RBPs associated with ALS-specific fusions in ALS (*left*) and C9-ALS (*right*). Dot size represents statistical significance measured as - log_10_(FDR), while color intensity reflects the magnitude of expression change measured as log_2_(Fold Change). Upregulated RBPs are shown in red, and downregulated RBPs are in blue. Gray indicates RBPs that are not significant DEGs, while absence of a dot denotes lack of expression in the corresponding tissue. **(b)** Individual-value plot demonstrating increased ELAVL2 protein levels in ALS mCTX (n = 19) compared to controls (n = 16) (Mann-Whitney U test = 88, p = 0.0343). **(c)** Individual-value plot revealing higher ELAVL2 protein levels in C9-ALS mCTX (n = 12) compared to controls (n = 16) (Mann-Whitney U test = 47, p = 0.0226). **(d)** Correlation scatter plot showing the relationship between ELAVL2 protein levels and disease duration in ALS mCTX. Higher ELAVL2 levels were significantly associated with shorter disease duration (Spearman r = -0.5223, p = 0.0218). **(e)** Correlation scatter plot demonstrating the relationship between ELAVL2 protein levels and disease duration in C9-ALS mCTX. No significant correlation was observed (Spearman r = -0.1763, p = 0.6238). **(f)** LOGO plot showing the motif pattern of *ELAVL2*. The *E*-value above the plot (*E* = 5.27e-5) indicates the significance of motif enrichment in the 200 bp region surrounding fusion junctions compared to the 200 bp surrounding canonical splice sites. The ratios in brackets (91.1%/76.3%) show the motif frequency at all ALS-specific fusion sites (*left*) and at all canonical splice sites (*right*). **(g)** Curve plot showing the positional density of *ELAVL2* motifs across a 200 bp window surrounding the fusion junction (red curve along the *top* cartoon on the x-axis) or the canonical splice sites (blue curve along the *bottom* cartoon on the x-axis). * p < 0.05; ** p < 0.01.

To further validate dysregulation of fusion-associated RBPs, we used ELAVL2 as an example and first assessed protein levels by western blot. ELAVL2 levels were significantly increased in ALS (Fig. 6b) and C9-ALS mCTX (Fig. 6c) compared to controls. ELAVL2 levels were also significantly higher in males but not females in both ALS (Suppl. Fig. 7b and c) and C9-ALS (Suppl. Fig. 7d and e). Lastly, in ALS, ELAVL2 levels were inversely correlated with disease duration, with higher ELAVL2 associated with shorter disease duration (Fig. 6d). No significant correlation was observed in C9-ALS (Fig. 6e).

Additionally, 10 of the 12 RBPs (except *HNRNPR* and *RBMS3*) are involved in RNA 3’ end cleavage and polyadenylation.^35,53–62^ Given that the identified ALS-specific fusion junctions were not always at the 3’ end/ 3’ untranslated region (UTR) of the parent genes, we further compared the locations of these motifs around fusion junctions to those around canonical splice junctions. The largest differences were observed at 50bp upstream of both junction sites, particularly at the left junction. The left fusion junctions had a total depletion of all these motifs, whereas the canonical left splice junctions had peaked motif frequencies (Fig. 6f and g, Supplementary Fig. 8). These findings suggest that the relationship between these RBPs and ALS-specific fusion may be more complex and involve novel mechanisms.

#### ALS-specific RNA fusions are associated with GTPase-mediated pathways

Although individual RNA fusion events are rare, their cumulative burden in ALS may be substantial. Therefore, we hypothesized that ALS-specific RNA fusions could be associated with transcriptomic dysregulation that was previously overlooked. This hypothesis also implies that accounting for the presence of ALS-specific RNA fusion might clarify the unexplained variance in our regression models for transcriptional disruptions. None of the 17,679 GE outliers between ALS and control samples was related to the occurrence of ALS-specific RNA fusions. In contrast, 28 of 301 mis-splicing outliers were significantly associated with ALS-specific fusions (Fig. 7a, Supplementary Table 8). GO enrichment identified an enrichment for GTPase-related and ligand-gated activities (Fig. 7b). These patterns were absent among 16 mis-splicing outliers associated with control-specific fusions (Supplementary Fig. 9a and b) and among all 301 mis-splicing outliers regardless of fusion (Supplementary Fig. 9c), indicating that GTPase-related and ligand-gated pathway enrichment was specific to mis-splicing outliers associated with ALS-specific fusions.

**Figure 7.**
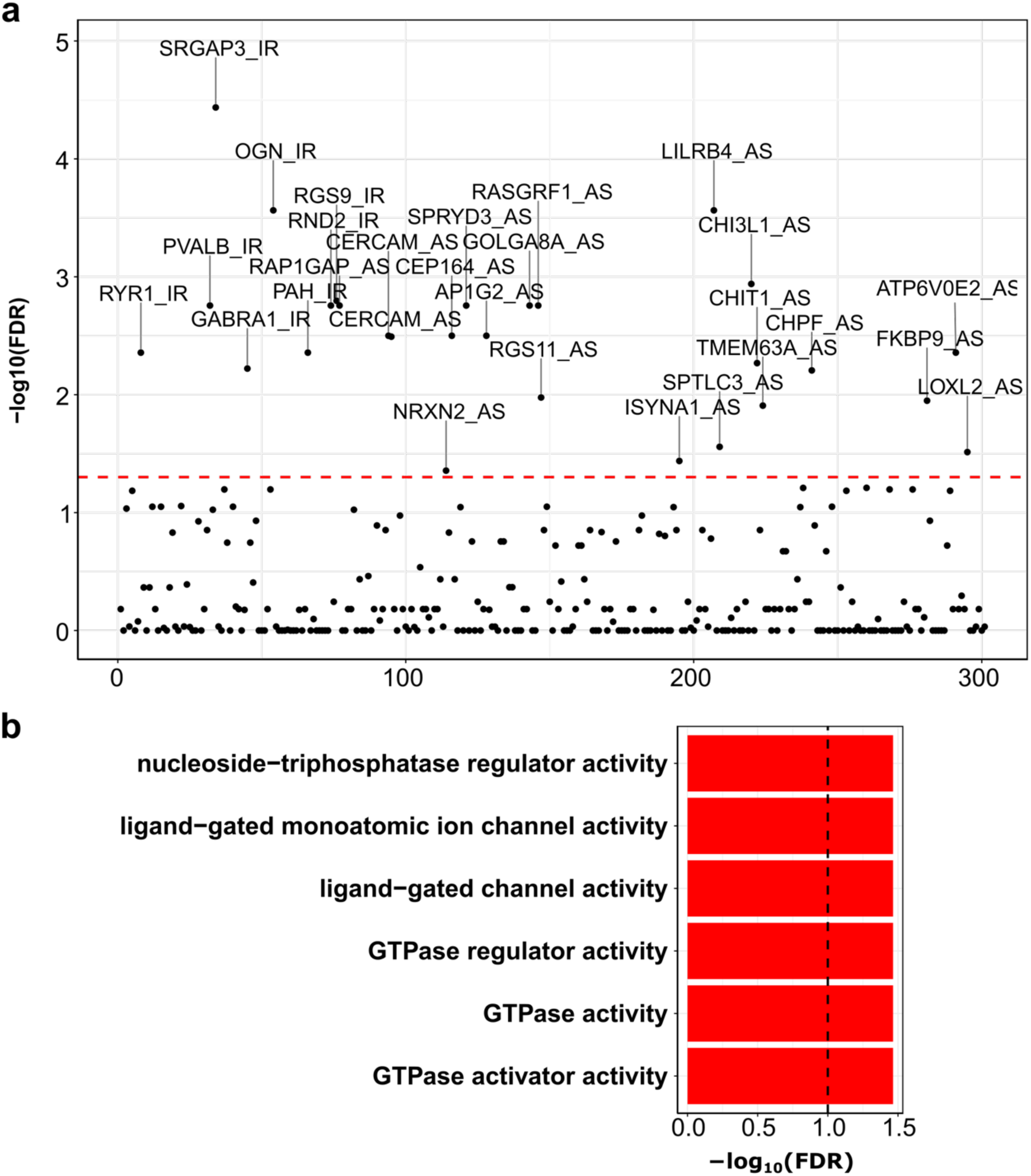
ALS-specific fusions may exacerbate ALS pathology by impacting specific pathways. **(a)** Scatter plot identifying splicing outliers associated with ALS-specific fusions. Each dot represents a splicing outlier event, annotated as either AS or IR; the x-axis denotes the outlier index, and the y-axis indicates the statistical significance of the association measured as - log_10_(FDR). The red horizontal dashed line represents the cutoff of significance. **(b)** GO enrichment analysis of host genes harboring splicing outliers associated with ALS-specific fusions. The x-axis represents the enrichment significance measured as -log_10_(FDR), and the y-axis shows GO terms.

## Discussion

In this study, we provide an integrated view of transcriptomic dysregulation in ALS by combining a reference-guided framework for GE and splicing with *de novo* discovery of fusion transcripts across five post-mortem CNS tissues. By modelling DEGs together with DAS and DIR, and extending beyond annotation to capture chimeric RNA events, we advance a unified concept of transcriptomic instability in ALS, where disrupted RNA processing is accompanied by a heightened burden of non-canonical transcriptional events. Together, these analyses reveal that RNA-processing defects represent a dominant axis of transcriptomic alteration in ALS.

Mis-splicing emerged as the dominant disease signal in our reference-guided analyses, indicating a widespread loss of splicing fidelity across ALS rather than differential gene expression.^8^ Within this landscape, AS was extensively dysregulated across ALS CNS tissues, particularly in neurons as demonstrated previously.^8,63^ Furthermore, widespread exon mis-regulation in ALS was strongly linked to loss of function of key RBPs similar to previous findings where most notably TDP-43 has been shown to induce aberrant splicing and cryptic exon inclusion.^64–67^

In our analyses, IR represented the most prevalent and quantitatively dominant form of mis-splicing and the dominant transcriptomic abnormality overall. This observation aligns with evidence that IR is a reproducible and biologically meaningful component of RNA dysregulation in ALS rather than a generic artefact of degeneration and has been documented in both cellular models and patient tissues, including C9-ALS.^68–71^

Several non-mutually exclusive mechanisms could explain why ALS becomes “IR-prone”. Neuronal aging and chronic stress can destabilize RBP and spliceosome homeostasis, while mislocalization of splicing factors, such as TDP-43 and FUS, may impair RNA surveillance and promote accumulation of aberrant intron-containing transcripts.^72–74^ IR may also exhibit cell-type-specific regulation, as altered intronic processing has been reported in reactive astrocytes.^75^

Our integrative pathway analyses underscore both divergence and convergence across tissues and genotypes. Immune and neuroinflammatory signatures were prominent, consistent with extensive evidence implicating microglia and other glial cells in ALS pathogenesis.^76^ In parallel, non-immune pathways, including phospholipid metabolism,^77^ neurite development, and cytoskeletal organization,^78^ emerged across tissues. Importantly, immune enrichment was largely driven by expression-level changes, whereas mis-splicing preferentially affected neuronal and intracellular pathways, highlighting a functional dissociation. We also observed a consistent convergence on small GTPase signaling across tissues and ALS subtypes consistent with previous evidence implicating multiple GTPase pathways in ALS^79–87^ and the intersection of C9ORF72 biology with GTPase regulation.^88–91^

Previous bulk transcriptome analyses have reported distinct patterns in C9-ALS compared to ALS across brain regions.^20^ Our results place these differences within a unifying framework: ALS and C9-ALS share a transcriptomic architecture dominated by mis-splicing but differ in downstream pathway engagement. Immune-related pathways were enriched in spinal cord of C9-ALS, whereas neuronal and structural pathways were more prominently disrupted in ALS. Splicing factors and RBPs were extensively disrupted in both groups, although C9-ALS exhibited a milder or more downregulated splicing profile. These differences were also reflected in cellular signatures. ALS and C9-ALS shared enrichment of microglial gene expression in spinal cord,^14^ whereas C9-ALS exhibited enrichment of excitatory neuron markers in FC, consistent with region-specific neuronal state perturbation reported in ALS and C9-ALS.^44,92,93^ Finally, sex differences are also well-recognized in ALS, with higher incidence and more aggressive clinical course in men.^46,47,94^ Consistently, our analyses revealed sex as an additional source of transcriptomic heterogeneity, with male-associated effects intersected with neurite and cytoskeletal biology and broader pathway disruption. Together, these findings indicate that genotype, cellular context, and sex collectively shape the transcriptomic heterogeneity of ALS.

Our *de novo* analyses reveal an increased burden of fusion transcripts in ALS, extending evidence that chimeric transcription is detectable in ALS transcriptomes.^21,22^ Recapitulation of ALS-specific fusion transcripts in an independent cohort using PacBio long-read sequencing, together with junction-specific BaseScope detection of *FAM69A-EVI5* in ALS cerebellum, provides orthogonal support for the biological existence of these fusions beyond short-read sequencing. Although individual fusions are likely restricted to subsets of individuals living with ALS, our findings support a model in which impaired RNA processing generates heterogeneous fusion transcripts that may represent molecular signature of disrupted RNA-processing fidelity.

Unlike cancer-associated fusions, which are mainly driven by genomic rearrangements,^50^ our integration of WGS and RNASeq supports ALS-specific fusions as RNA-only events arising independently of SVs. This is consistent with previous work in ALS iPSC-derived motor neurons reporting fusions associated with splicing alterations and TDP-43 proteinopathy.^21^ Our motif analyses further support altered splice-site definition at ALS-specific fusion junctions, involving RBPs,^58,59^ most of which participate in RNA 3’ cleavage and polyadenylation, processes increasingly implicated in ALS transcriptomic dysregulation.^95,96^ ELAVL2 provided an example of this broader fusion-associated RBP dysregulation in human tissue, with increased protein levels in both ALS and C9-ALS mCTX which correlated with shorter disease duration in ALS.

Fusion-like chimeric RNAs are not unique to disease and have been reported to increase with age, suggesting that baseline fusion formation may reflect physiological transcriptional plasticity or age-associated transcriptional noise.^97^ The excess fusion burden observed in ALS may reflect amplification of this baseline process in the context of impaired RNA processing. ALS-specific fusion occurrence was also associated with severe splicing outliers in genes governing GTPase activity, suggesting convergence between quantitative mis-splicing and qualitative fusion events on shared pathways.

This study has several limitations. Post-mortem tissue captures a mixture of causal, compensatory, and terminal changes, and residual confounding by agonal state, RNA integrity, and treatment history cannot be fully excluded. Bulk tissue analyses cannot definitively assign mis-splicing or fusion events to specific cell types, and deconvolution cannot replace direct single-cell measurement of RNA processing. Fusion detection from short-read RNASeq remains inherently error-prone, and even conservative filtering cannot entirely exclude artefacts. Although long-read RNASeq provided independent confirmation of multiple fusion events and junction-specific BaseScope supported the endogenous presence of *FAM69A-EVI5* in ALS tissue, these validations were applied to selected events and do not exclude false positives among the remaining fusion calls, particularly given the smaller number of non-neurological controls. Finally, the observational nature of human post-mortem data limits causal inference, and it remains unclear whether IR and fusion burden represent early drivers, late consequences, or both.

In summary, this study redefines the ALS transcriptome as a landscape of structural RNA disruption. From the global accumulation of retained introns to the generation of novel, non-genomic fusions, the disease is characterized by a collapse in processing fidelity associated with disruption of GTPase-related pathways. Functional studies will be required to determine the consequences of ALS-specific fusions and whether these events actively contribute to disease mechanisms. Together, our findings highlight the importance of extending expression-based approaches to include splicing-centric and RNA-structural therapeutic strategies.

## Data availability

The short-read RNASeq and WGS datasets of the NYGC cohort are available via the consortia portal (https://dataengine.targetals.org/collections/postmortem-tissue-core/overview)

The long-read PacBio Iso-Seq dataset of the MGH cohort is available in dbGaP.

## Author contributions

HX, TP and AB contributed to study design, data collection, data analysis and drafting of the manuscript. SS, SSH, AS, XZ, MK, EJG, ALCT, RZBM, JL, BJ, and CEFDE contributed to data collection, data analysis, and editing of the manuscript. MEC, JDB, HB, MET and RMP contributed to study design and editing of the manuscript. DG and GSV contributed to study design, supervised the study and drafted the manuscript.

## Acknowledgements

The authors would like to thank the Target ALS Human Postmortem Tissue Core, New York Genome Center for Genomics of Neurodegenerative Disease, Amyotrophic Lateral Sclerosis Association and TOW Foundation. All NYGC ALS Consortium activities are supported by the ALS Association (ALSA, 19-Si-459) and the Tow Foundation. The graphical abstract was created with BioRender.com.

## Supplementary material

**Appendix 1**

NYGC ALS Consortium

Hemali Phatnani^5^, Justin Kwan^6^, Dhruv Sareen^7,8^, James R. Broach^9^, Zachary Simmons^10^, Ximena Arcila-Londono^11^, Edward B. Lee^12^, Vivianna M. Van Deerlin^12^, Neil A. Shneider^13^, Ernest Fraenkel^1^, Lyle W. Ostrow^14^, Frank Baas^15,16^, Noah Zaitlen^17^, James D. Berry^18,19^, Andrea Malaspina^19,20,21^, Pietro Fratta^22^ , Gregory A. Cox^23^, Leslie M. Thompson^24,25^, Steve Finkbeiner^26^, Efthimios Dardiotis^27^, Timothy M. Miller^28^, Siddharthan Chandran^29^, Suvankar Pal^29^, Eran Hornstein^30^, Daniel J. MacGowan^31^, Terry Heiman-Patterson^32^, Molly G. Hammell^33^, Nikolaos. A. Patsopoulos^34,35^, Oleg Butovsky^36^, Joshua Dubnau^37^, Avindra Nath^38^, Robert Bowser^39,40^, Matthew Harms^41^, Eleonora Aronica^42^, Mary Poss^43^, Jennifer Phillips-Cremins^44^, John Crary^45^, Nazem Atassi^46^, Dale J. Lange^47,48^, Darius J. Adams^49,50^, Leonidas Stefanis^51,52^, Marc Gotkine^53^, Robert H. Baloh^54,55^, Suma Babu^19^, Towfique Raj^56^, Sabrina Paganoni^57^, Ophir Shalem^58,59^, Colin Smith^60,61^, Bin Zhang^62^, Brent Harris^63^, Iris Broce^64^, Vivian Drory^65^, John Ravits^66^, Corey McMillan^67^, Vilas Menon^68^, Lani Wu^69^, Steven Altschuler^69^, Yossef Lerner^70^, Rita Sattler^71^, Kendall Van Keuren-Jensen^72^, Orit Rozenblatt-Rosen^73^, Kerstin Lindblad-Toh^73^, Katharine Nicholson^74^, Peter Gregersen^75^, Jeong-Ho Lee^76^, Sulev Kokos^77^, Stephen Muljo^78^, & Bryan J. Traynor^79^.

^5^Center for Genomics of Neurodegenerative Disease (CGND), New York Genome Center, New York, NY, USA. ^6^Department of Neurology, Lewis Katz School of Medicine, Temple University, Philadelphia, PA, USA. ^7^Cedars-Sinai Department of Biomedical Sciences, Board of Governors Regenerative Medicine Institute and Brain Program, Cedars-Sinai Medical Center, University of California, Los Angeles, CA, USA. ^8^Department of Medicine, University of California, Los Angeles, CA, USA. ^9^Department of Biochemistry and Molecular Biology, Penn State Institute for Personalized Medicine, The Pennsylvania State University, Hershey, PA, USA. ^10^Department of Neurology, The Pennsylvania State University, Hershey, PA, USA. ^11^Department of Neurology, Henry Ford Health System, Detroit, MI, USA. ^12^Department of Pathology and Laboratory Medicine, Perelman School of Medicine, University of Pennsylvania, Philadelphia, PA, USA. ^13^Department of Neurology, Center for Motor Neuron Biology and Disease, Institute for Genomic Medicine, Columbia University, New York, NY, USA. ^1^Department of Biological Engineering, Massachusetts Institute of Technology, Cambridge, MA, USA. ^14^Department of Neurology, Johns Hopkins School of Medicine, Baltimore, MD, USA. ^15^Department of Neurogenetics, Academic Medical Centre, Amsterdam, The Netherlands. ^16^Leiden University Medical Center, Leiden, The Netherlands. ^17^Department of Medicine, Lung Biology Center, University of California, San Francisco, CA, USA. ^18^ALS Multidisciplinary Clinic, Neuromuscular Division, Department of Neurology, Harvard Medical School, Boston, MA, USA. ^19^Neurological Clinical Research Institute, Massachusetts General Hospital, Boston, MA, USA. ^19^Centre for Neuroscience and Trauma, Blizard Institute, Barts, Queen Mary University of London, London, UK. ^20^The London School of Medicine and Dentistry, Queen Mary University of London, London, UK. ^21^Department of Neurology, Basildon University Hospital, Basildon, UK. ^22^Institute of Neurology, National Hospital for Neurology and Neurosurgery, University College London, London, UK. ^23^The Jackson Laboratory, Bar Harbor, ME, USA. ^24^Department of Psychiatry and Human Behavior, Department of Biological Chemistry, School of Medicine, University of California, Irvine, CA, USA. ^25^Department of Neurobiology and Behavior, School of Biological Sciences, University of California, Irvine, CA, USA. ^26^Taube/Koret Center for Neurodegenerative Disease Research, Roddenberry Center for Stem Cell Biology and Medicine, Gladstone Institute, San Francisco, CA, USA. ^27^Department of Neurology and Sensory Organs, University of Thessaly, Thessaly, Greece. ^28^Department of Neurology, Washington University in St Louis, St Louis, MO, USA. ^29^Centre for Clinical Brain Sciences, Anne Rowling Regenerative Neurology Clinic, Euan MacDonald Centre for Motor Neurone Disease Research, University of Edinburgh, Edinburgh, UK. ^30^Department of Molecular Genetics, Weizmann Institute of Science, Rehovot, Israel. ^31^Department of Neurology, Icahn School of Medicine at Mount Sinai, New York, NY, USA. ^32^Center for Neurodegenerative Disorders, Department of Neurology, the Lewis Katz School of Medicine, Temple University, Philadelphia, PA, USA. ^33^Cold Spring Harbor Laboratory, Cold Spring Harbor, NY, USA. ^34^Computer Science and Systems Biology Program, Ann Romney Center for Neurological Diseases, Department of Neurology and Division of Genetics in Department of Medicine, Brigham and Women’s Hospital, Harvard Medical School, Boston, MA, USA. ^35^Program in Medical and Population Genetics, Broad Institute, Cambridge, MA, USA. ^36^Ann Romney Center for Neurologic Diseases, Brigham and Women’s Hospital, Harvard Medical School, Boston, MA, USA. ^37^Department of Anesthesiology, Stony Brook University, Stony Brook, NY, USA. ^38^Section of Infections of the Nervous System, National Institute of Neurological Disorders and Stroke, NIH, Bethesda, MD, USA. ^39^Department of Neurology, Barrow Neurological Institute, St Joseph’s Hospital, Phoenix, AZ, USA. ^40^Medical Center, Department of Neurobiology, Barrow Neurological Institute, St Joseph’s Hospital and Medical Center, Phoenix, AZ, USA. ^41^Department of Neurology, Division of Neuromuscular Medicine, Columbia University, New York, NY, USA. ^42^Department of Neuropathology, Academic Medical Center, University of Amsterdam, Amsterdam, The Netherlands. ^43^Department of Biology and Veterinary and Biomedical Sciences, The Pennsylvania State University, University Park, PA, USA. ^44^New York Stem Cell Foundation, Department of Bioengineering, School of Engineering and Applied Sciences, University of Pennsylvania, Philadelphia, PA, USA. ^45^Department of Pathology, Fishberg Department of Neuroscience, Friedman Brain Institute, Ronald M. Loeb Center for Alzheimer’s Disease, Icahn School of Medicine at Mount Sinai, New York, NY, USA. ^46^Department of Neurology, Harvard Medical School, Neurological Clinical Research Institute, Massachusetts General Hospital, Boston, MA, USA. ^47^Department of Neurology, Hospital for Special Surgery, New York, NY, USA. ^48^Weill Cornell Medical Center, New York, NY, USA. ^49^Medical Genetics, Atlantic Health System, Morristown Medical Center, Morristown, NJ, USA. ^50^Overlook Medical Center, Summit, NJ, USA. ^51^Center of Clinical Research, Experimental Surgery and Translational Research, Biomedical Research Foundation of the Academy of Athens (BRFAA), Athens, Greece. ^52^1st Department of Neurology, Eginition Hospital, Medical School, National and Kapodistrian University of Athens, Athens, Greece. ^53^Neuromuscular/EMG service and ALS/Motor Neuron Disease Clinic, Hebrew University-Hadassah Medical Center, Jerusalem, Israel. ^54^Board of Governors Regenerative Medicine Institute, Los Angeles, CA, USA. ^55^Department of Neurology, Cedars-Sinai Medical Center, Los Angeles, CA, USA. ^56^Departments of Neuroscience, and Genetics and Genomic Sciences, Ronald M. Loeb Center for Alzheimer’s disease, Icahn School of Medicine at Mount Sinai, New York, NY, USA. ^57^Harvard Medical School, Department of Physical Medicine and Rehabilitation, Spaulding Rehabilitation Hospital, Boston, MA, USA. ^58^Center for Cellular and Molecular Therapeutics, Children’s Hospital of Philadelphia, Philadelphia, PA, USA. ^59^Department of Genetics, Perelman School of Medicine, University of Pennsylvania, Philadelphia, PA, USA. ^60^Centre for Clinical Brain Sciences, University of Edinburgh, Edinburgh, UK. ^61^Euan MacDonald Centre for Motor Neurone Disease Research, University of Edinburgh, Edinburgh, UK. ^62^Department of Genetics and Genomic Sciences, Icahn Institute of Data Science and Genomic Technology, Icahn School of Medicine at Mount Sinai, New York, NY, USA. ^63^Department of Neuropathology, Georgetown Brain Bank, Georgetown Lombardi Comprehensive Cancer Center, Georgetown University Medical Center, Washington, DC, USA. ^64^Neuroradiology Section, Department of Radiology and Biomedical Imaging, University of California, San Francisco, San Francisco, CA, USA. ^65^Neuromuscular Diseases Unit, Department of Neurology, Tel Aviv Sourasky Medical Center, Sackler Faculty of Medicine, Tel-Aviv University, Tel-Aviv, Israel. ^66^Department of Neuroscience, University of California San Diego, La Jolla, CA, USA. ^67^Department of Neurology, University of Pennsylvania Perelman School of Medicine, Philadelphia, PA, USA. ^68^Department of Neurology, Columbia University Medical Center, New York, NY, USA. ^69^Department of Pharmaceutical Chemistry, University of California San Francisco, San Francisco, CA, USA. ^70^Hadassah Hebrew University, Jerusalem, Israel. ^71^Department of Translational Neuroscience, Barrow Neurological Institute, Phoenix, AZ, USA. ^72^The Translational Genomics Research Institute (TGen), Phoenix, AZ, USA. ^73^Broad Institute, Cambridge, MA, USA. ^74^Massachusetts General Hospital, Boston, MA, USA. ^75^Institute of Molecular Medicine, Feinstein Institutes for Medical Research, Northwell Health, Manhasset, NY, USA. ^76^Korea Advanced Institute of Science and Technology (KAIST), Daejeon, South Korea. ^77^Perron Institute for Neurological and Translational Science, Nedlands, Western Australia, Australia. ^78^Integrative Immunobiology Section, National Institute of Allergy and Infectious Disease, NIH, Bethesda, MD, USA. ^79^Neuromuscular Disease Research Section, National Institute of Aging, NIH, Bethesda, MD, USA.

---

**Supplementary Figure 1.**
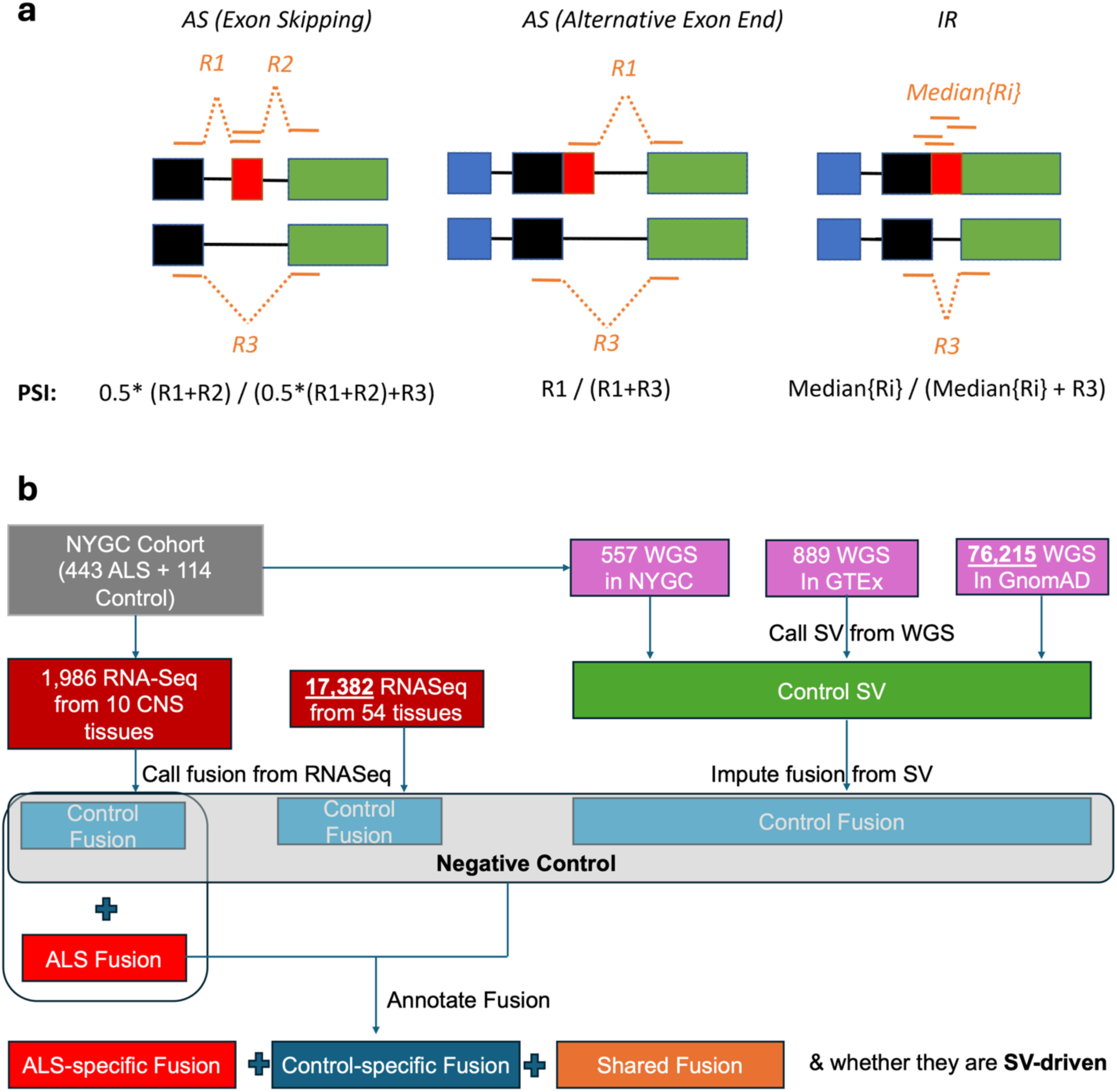
PSI calculation for mis-splicing and fusion benchmark. **(a)** Schematic representation and quantification of alternative splicing (AS), including “AS (Exon skipping)”, “AS (Alternative Exon End)” and intron retention (IR) events. R1, R2, and R3 represent RNASeq reads aligned to a given splice site, while Median {Ri} indicates the median count of reads aligned to the intronic region. **(b)** Schematic illustration of (1) assigning control-specific, ALS-specific, and shared to NYGC-RNASeq-derived fusion events and (2) benchmarking their potential of DNA rearrangement. (1) was done via querying a comprehensive source of SV annotations from the classification pipeline utilizing paired NYGC WGS, establishing GTEx RNASeq and gnomAD datasets as negative controls to distinguish fusion into three subtypes, namely ALS-specific fusion, control-specific fusion and shared fusion. This figure also refers to Fig. 4c.

**Supplementary Figure 2.**
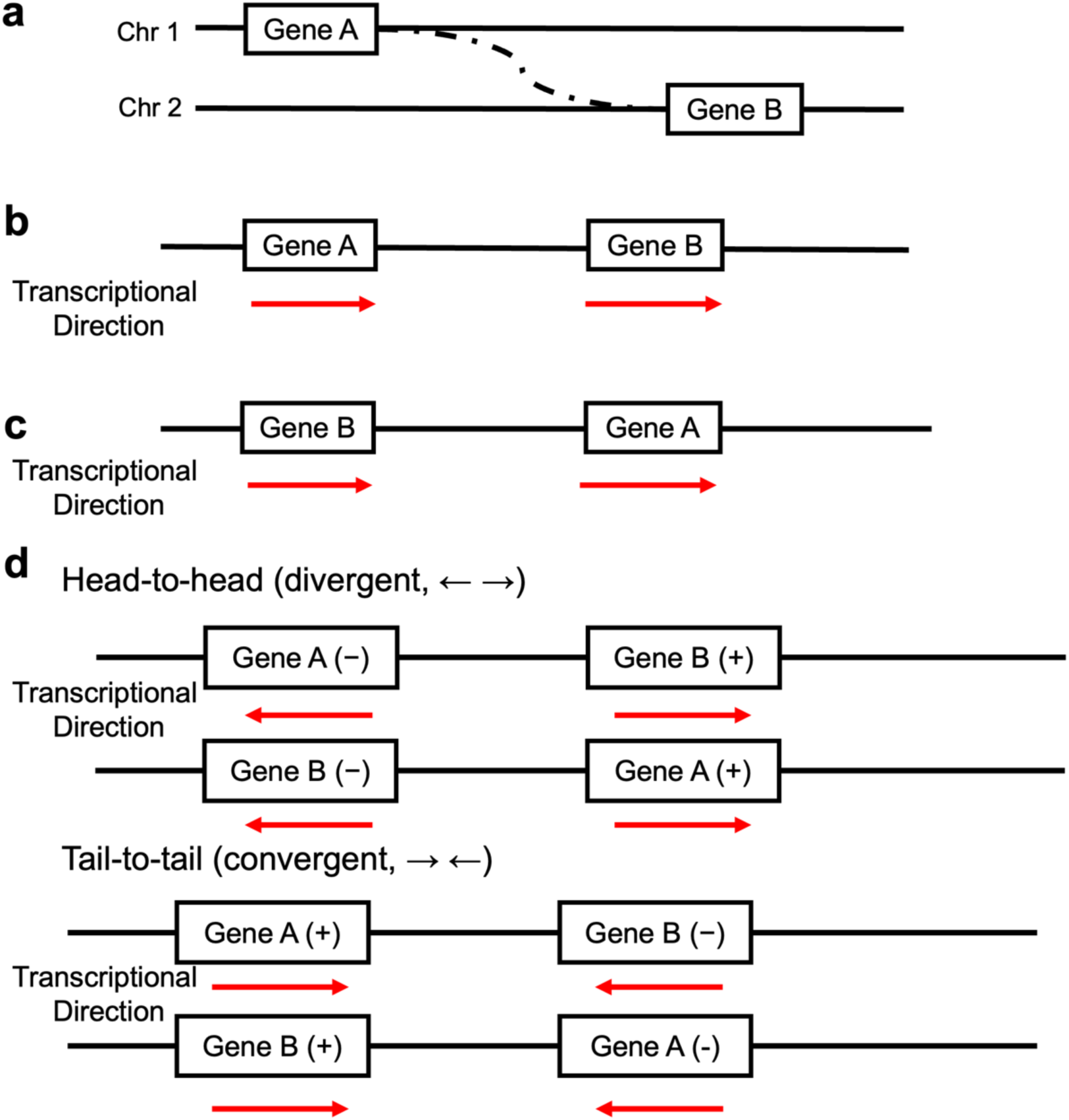
Categories of ALS-specific RNA fusions based on the chromosomal positions and strand orientations of the parent genes. Schematic representation of ALS-specific fusion categories defined by chromosomal origin and strand orientation of the parent genes. In all schemes, the observed fusion is “GeneA--GeneB” and the red arrows indicate the parent genes’ transcriptional directions. **(a)** Inter-chromosomal fusions, involving parent genes located on two different chromosomes. **(b)** Proximity fusions, involving genes on the same chromosome and strand; the upstream gene fuses with the downstream gene, concordant with their transcriptional directions. **(c)** Discordant orientation (same strand) fusions, involving genes on the same chromosome and strand; the downstream gene fuses with the upstream gene, in a discordant orientation relative to their transcriptional directions. **(d)** Discordant orientation (different strand) fusions, involving genes located on the same chromosome but opposite DNA strands; the fusion occurs in an inverted orientation relative to the reference genomic orchestration. This figure also refers to Fig. 4d.

**Supplementary Figure 3.**
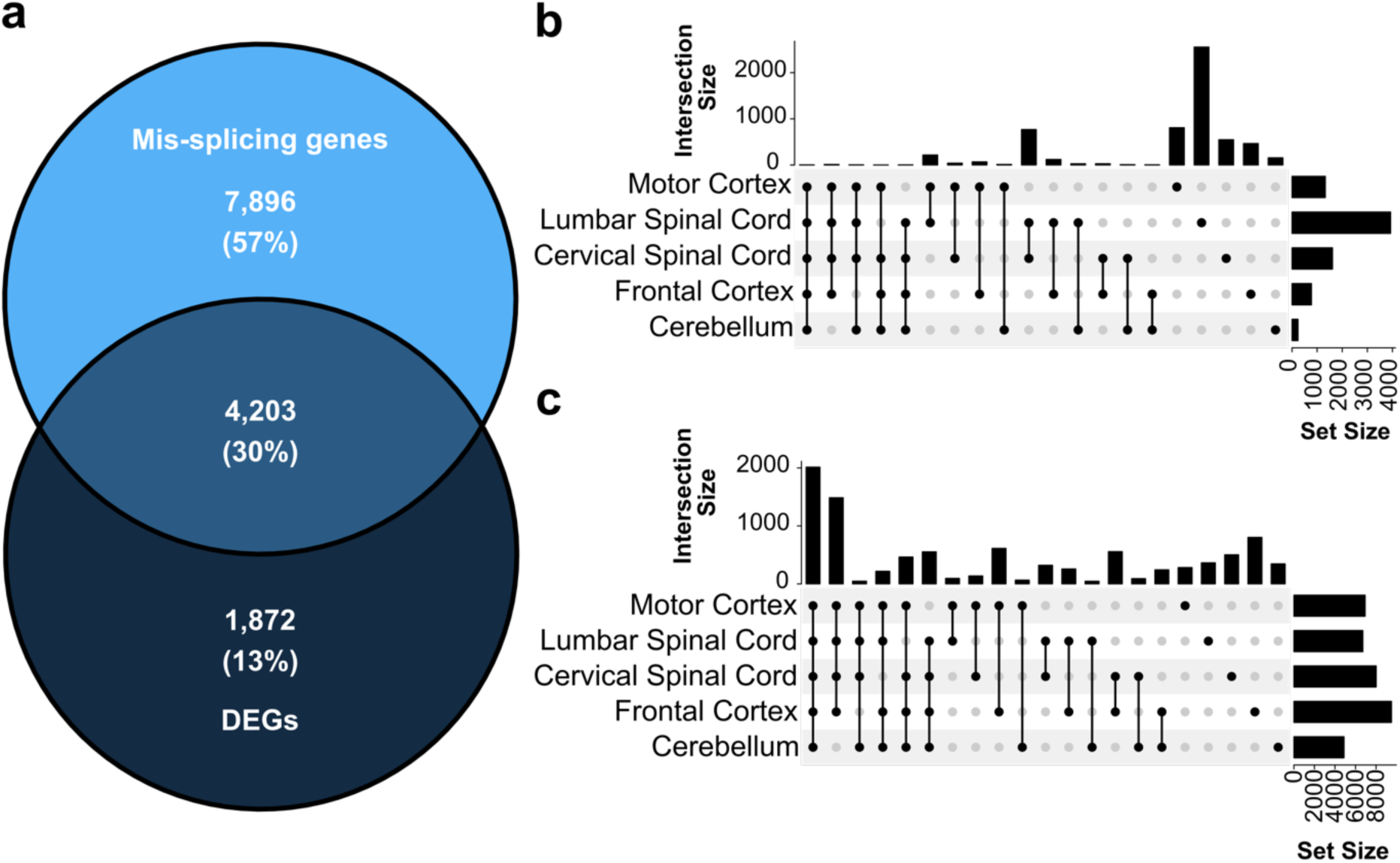
Breakdown of tissue-specific DEGs and mis-splicing. **(a)** Proportion of disrupted genes with mis-splicing (i.e. DIR or DAS; 57%) or differential expression (13%) in any of the five tissues. **(b)** UpSet plot shows the number of DEGs in each examined tissue. Horizontal bars represent the total count of DEGs in each tissue, while vertical bars represent the size of each intersection, corresponding to DEGs shared exclusively between specific tissues combinations. **(c)** UpSet plot shows the number of genes that are mis-spliced (i.e. DIR or DAS) in each tissue. Horizontal bars represent the total count of genes in each tissue, while vertical bars represent the size of each intersection, corresponding to genes shared exclusively between specific tissues combinations.

**Supplementary Figure 4.**
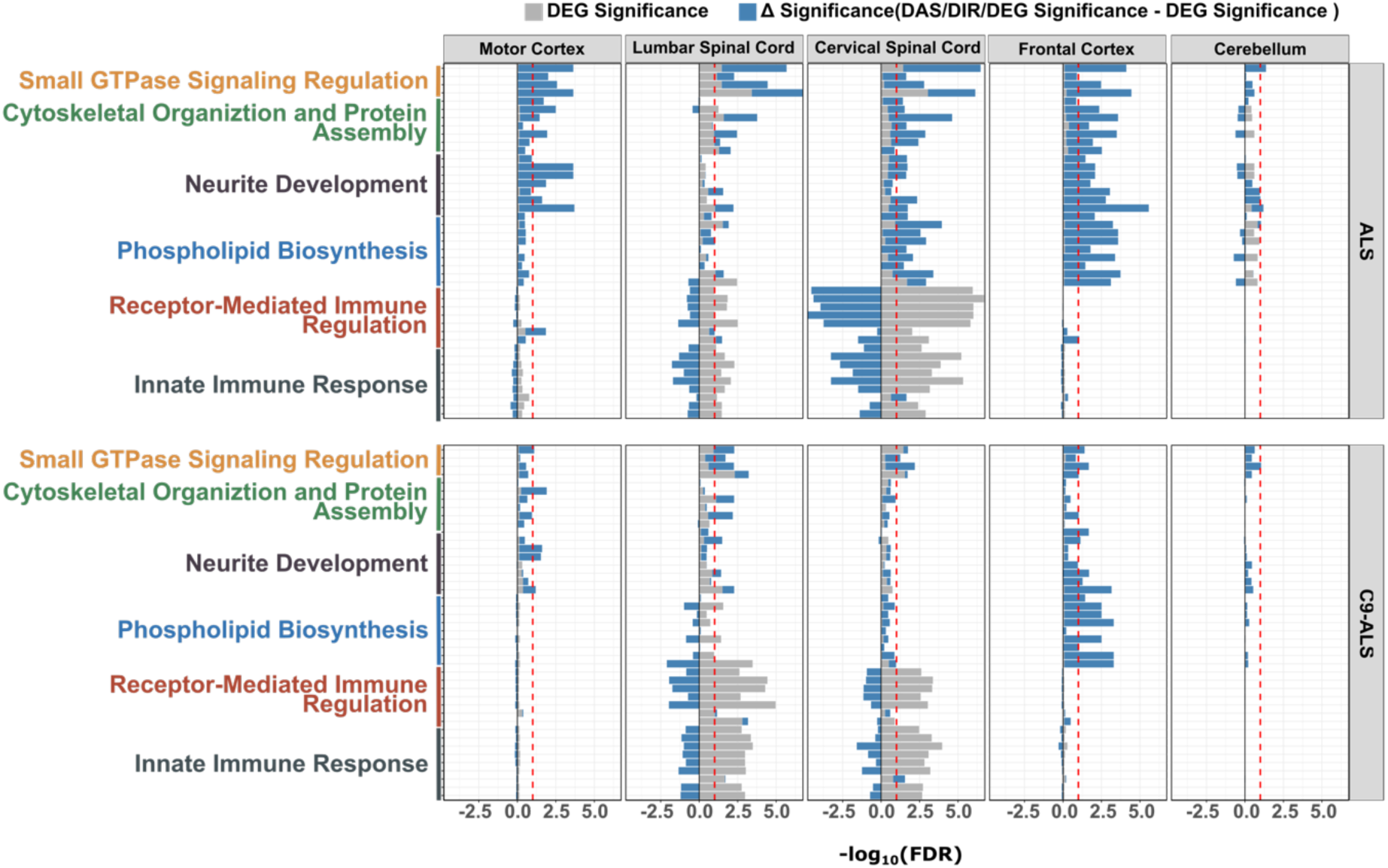
Aggregated functional enrichment of DEGs and mis-spliced genes unravels divergent biological pathways. The first round of GO enrichment significance, driven solely by DEGs and measured in - log_10_(FDR), is shown as grey bars. The red dashed line denotes the significance threshold for DEGs enrichment alone. The second round of GO enrichment was performed by adding host genes of mis-spliced events to DEGs, and the resulting changes in GO significance (ΔSignificance) are shown as blue bars. A positive blue bar indicates that mis-splicing amplifies functional disruption among DEGs, whereas a negative blue bar indicates that mis-splicing reduces it. The y-axis shows the same GO terms as Fig. 1d, although only the GO cluster names are showed. This figure also refers to Fig. 1d.

**Supplementary Figure 5.**
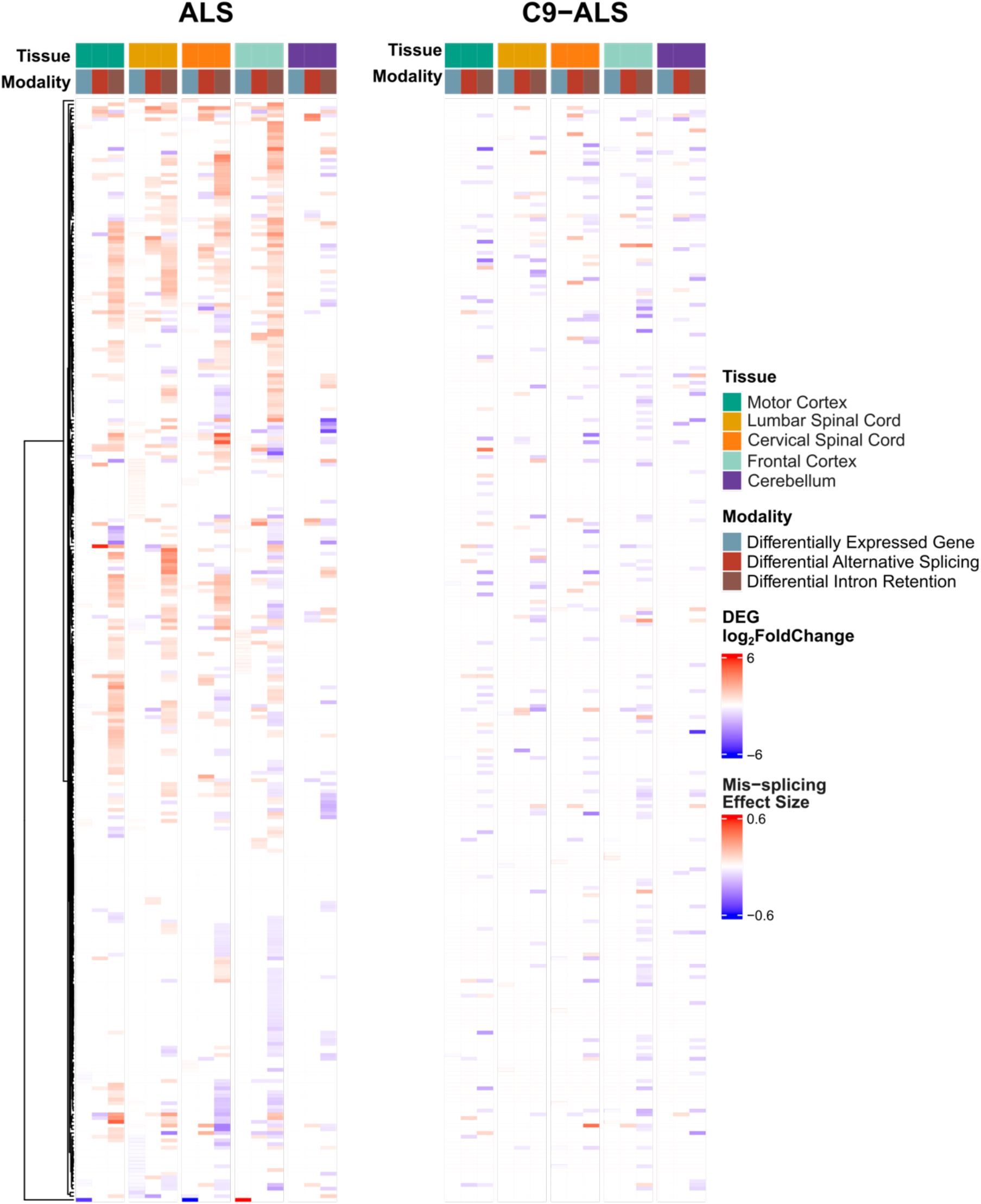
Full list of RBP enrichment analysis across ALS subtypes. The heatmap shows significant enrichment of transcriptional disruptions for RBPs and splicing factors in ALS (*left*) and C9-ALS (*right*). The top annotation bar includes two groups: the first row indicates tissue type (motor cortex in teal, lumbar spinal cord in yellow, cervical spinal cord in orange, frontal cortex in cyan and cerebellum in purple), while the second row denotes transcriptional modality (DEGs in grey, DASs in red, and DIRs in brown). Color intensity represents effect size for each ALS subtype compared to controls, with darker red indicating a larger effect size. For DEGs, effect size corresponds to the log_2_(Fold Change) in expression, whereas for DASs and DIRs, effect size corresponds to the maximum absolute ΔPSI value of all mis-spliced events within the gene. This figure also refers to Fig. 2a.

**Supplementary Figure 6.**
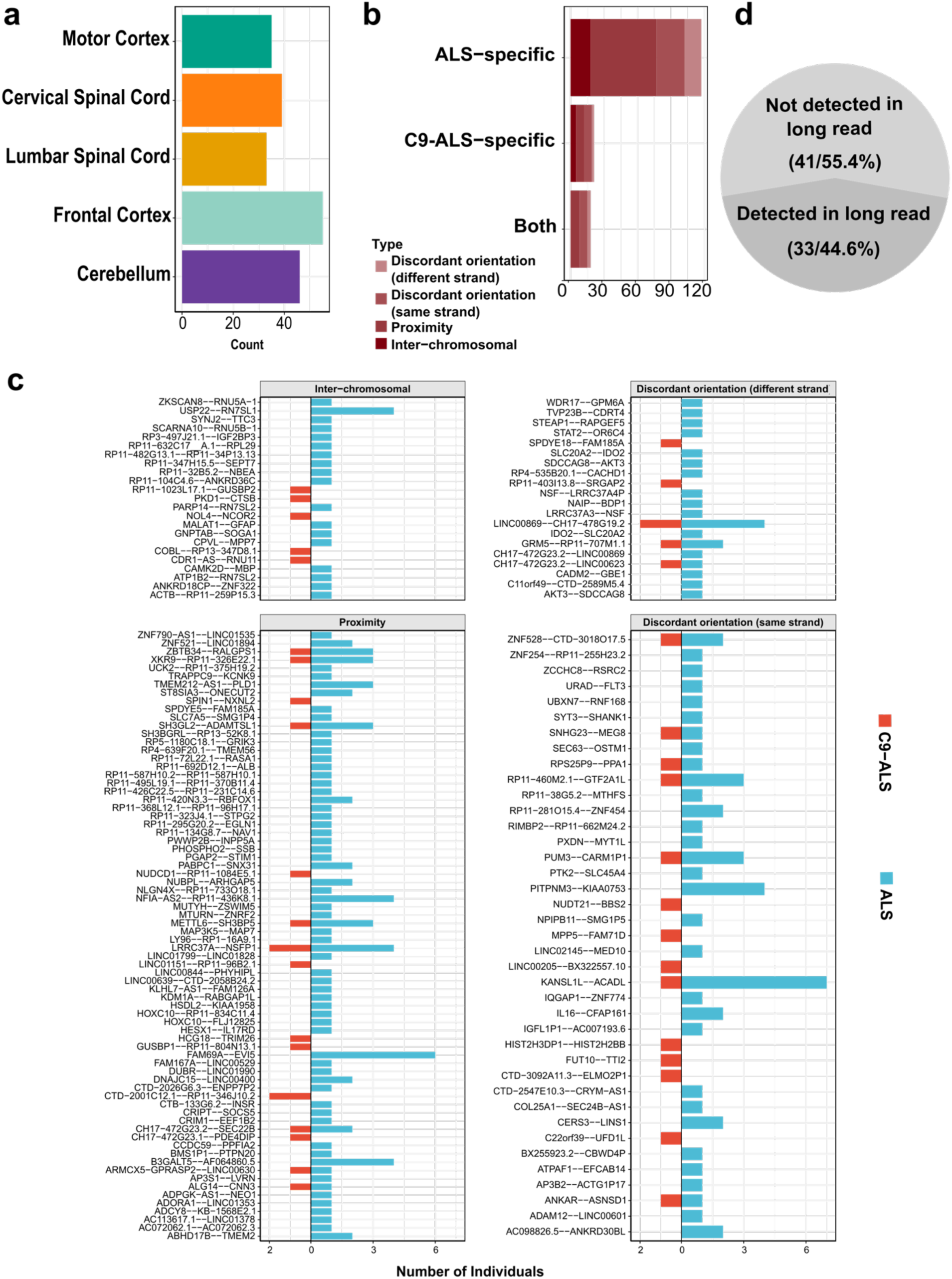
Distributions of ALS-specific RNA fusions across tissue and ALS subtypes. **(a)** Tissue-specific occurrence of ALS-specific fusions identified across five tissue types in the NYGC cohort. **(b)** Fusion occurrence per ALS subtypes (unique to ALS, unique to C9-ALS, or detected in both), with bar colors denoting the structural subtype as defined in Fig. 4d. **(c)** Number of ALS and C9-ALS individuals carrying each ALS-specific fusion in each fusion category. The x-axis represents the number of individuals, while the y-axis represents each ALS-specific fusion. Each section corresponds to an ALS-specific fusion category. This figure also refers to Fig. 4c-d.

**Supplementary Figure 7.**
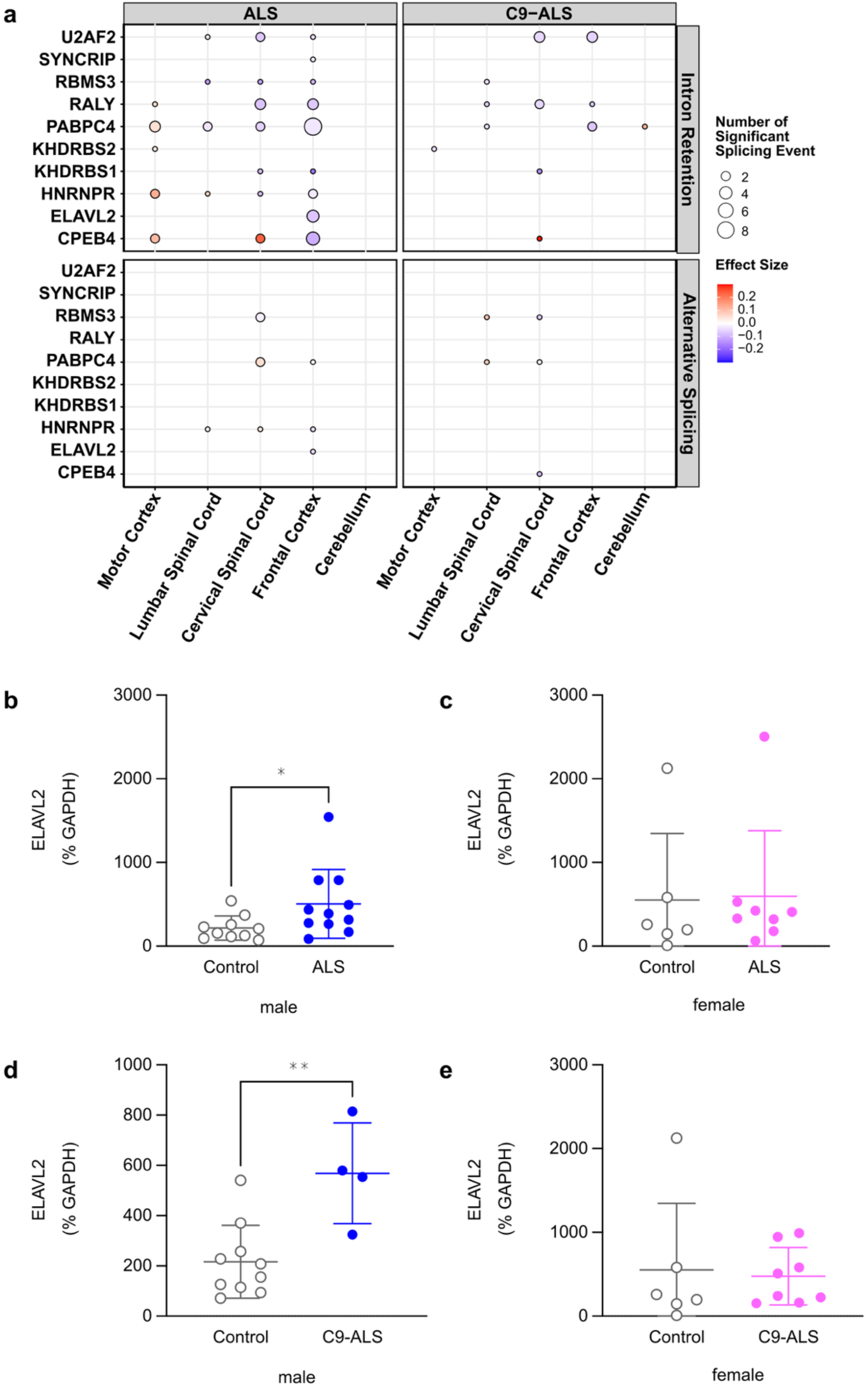
IR is dominant in the transcriptional disruption of RBPs associated with ALS-specific fusions. **(a)** Splicing dysregulation of RBPs associated with ALS-specific fusions in ALS (*left*) and C9-ALS (*right*). Dot size scales with the number of significant mis-splicing events (DAS or DIR) identified within each RBP, while color intensity represents the maximum effect size, measured in ΔPSI value, across these events (DAS or DIR). Red indicates exon inclusion or increased IR, whereas blue indicates exon skipping or decreased IR. This figure also refers to Fig. 5a. **(b)** Individual-value plot showing increased ELAVL2 protein levels in male ALS mCTX (n = 11) compared to male controls (n = 10) (Mann-Whitney U test = 23, p = 0.0242). **(c)** Individual-value plot revealing no significant differences in ELAVL2 protein levels between female ALS mCTX (n = 8) and female controls (n = 6) (Mann-Whitney U test = 19, p = 0.5728). **(d)** Individual-value plot demonstrating increased ELAVL2 protein levels in male C9-ALS mCTX (n = 4) compared to male controls (n = 10) (Mann-Whitney U test = 2, p = 0.0080). **(e)** Individual-value plot showing no significant differences in ELAVL2 protein levels between female C9-ALS mCTX (n = 8) and female controls (n = 6) (Mann-Whitney U test = 19, p = 0.5728). * p < 0.05; ** p < 0.01.

**Supplementary Figure 8.**
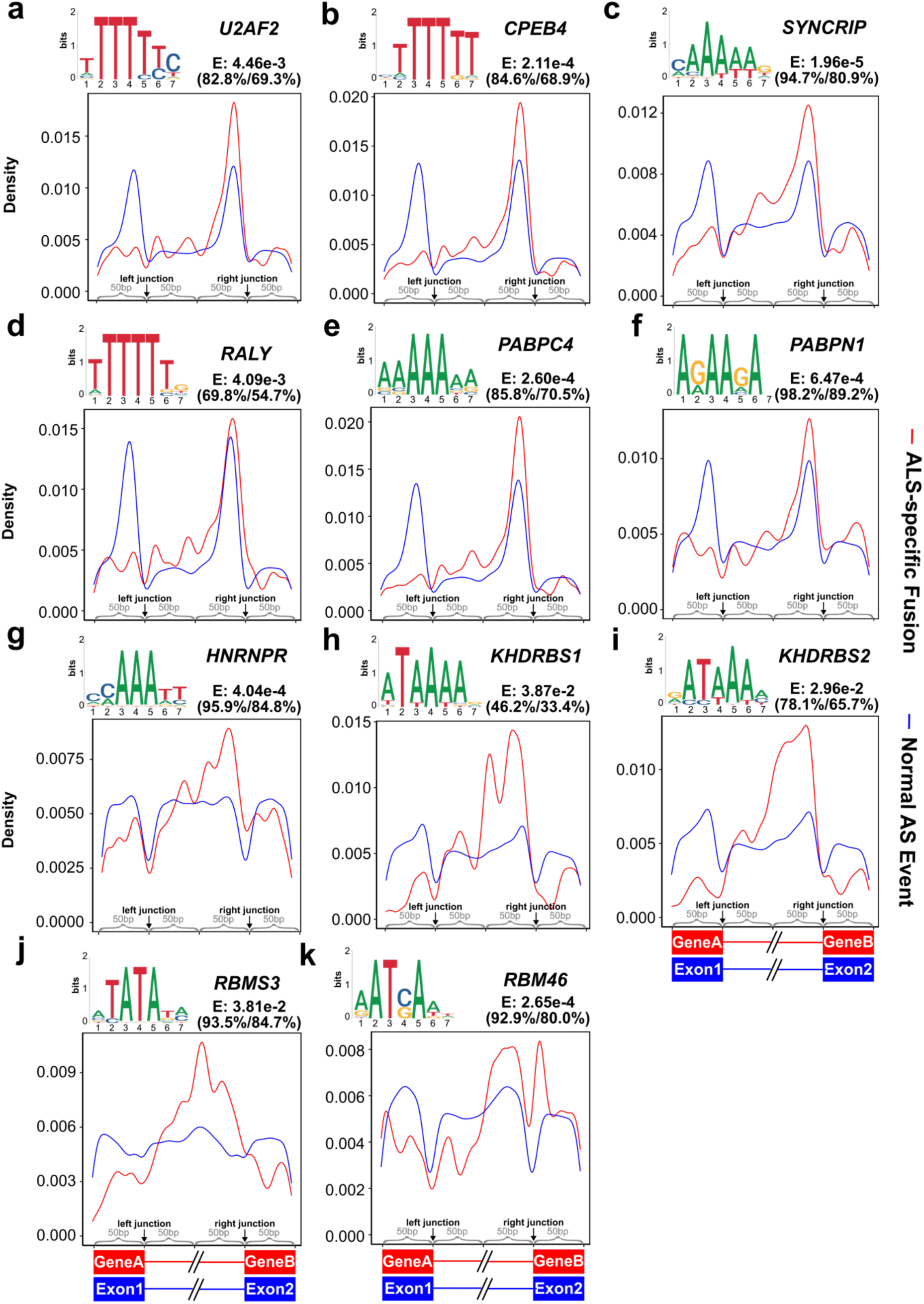
Motif positional patterns of RBPs enriched in ALS-specific fusions. **(a-k)** Motif patterns of all RBPs associated with ALS-specific fusion in LOGO plots and their positional densities across a 200 bp window surrounding the fusion junction (red line along the *top* cartoon of the x-axis) or the canonical splice sites (blue line along the *bottom* cartoon of the x-axis). Their enrichment *E*-values and relative frequencies are provided on the top of each LOGO plot. This figure also refers to Fig. 5b-c.

**Supplementary Figure 9.**
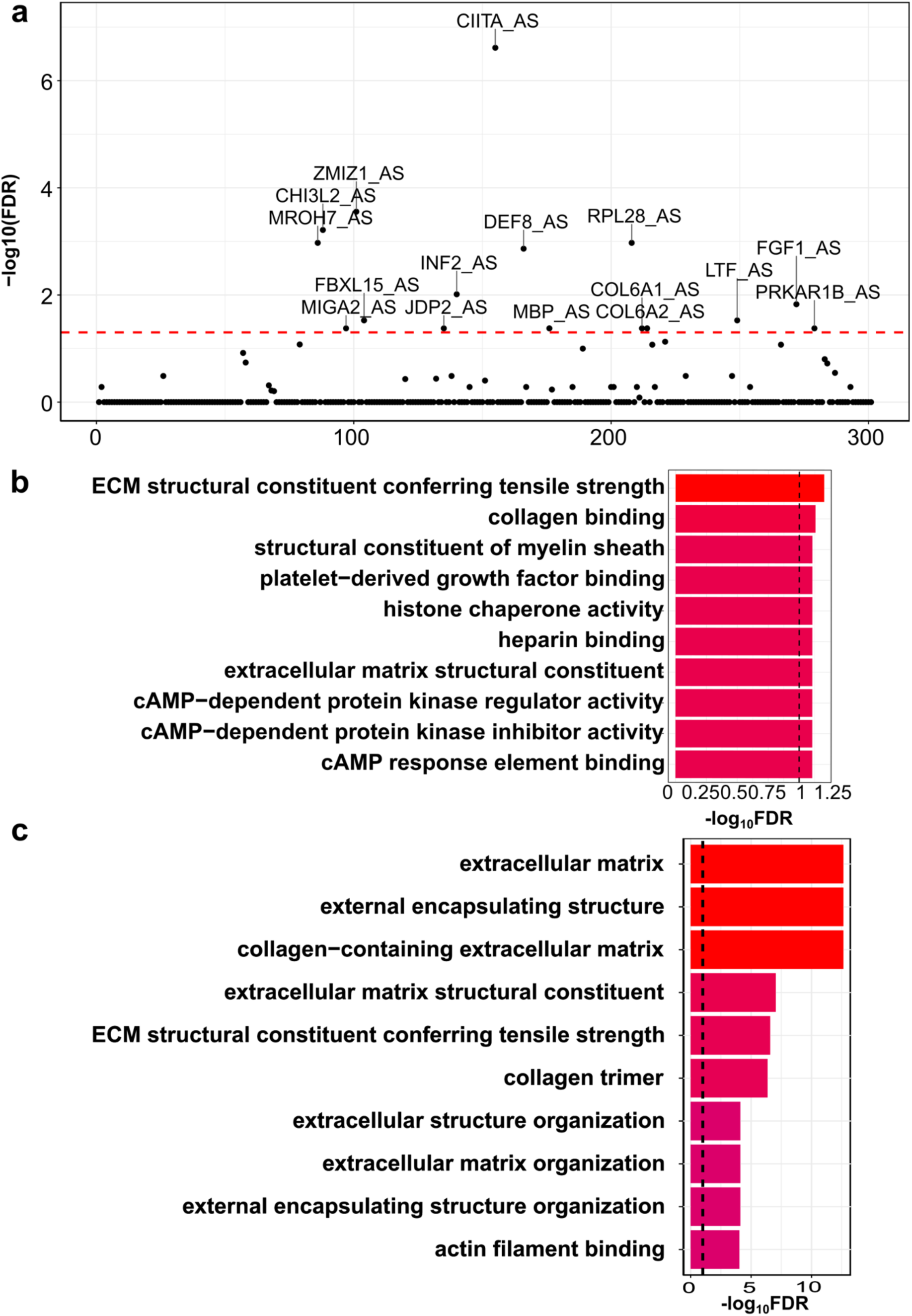
Functional enrichment of host genes harboring control-specific fusion associated splicing outliers. **(a)** Identification of splicing outliers associated with control-specific fusions. Each data point represents a unique splicing outlier event, annotated as either AS or IR. These events are the same events in Fig. 6a. The x-axis denotes the outlier index, and the y-axis indicates the statistical significance of the association measured in -log_10_(FDR). The red horizontal dashed line represents the cutoff of significance. **(b)** GO enrichment analysis of host genes harboring splicing outliers associated with control-specific fusions. The x-axis represents the enrichment significance measured in -log_10_(FDR), while the y-axis shows the GO terms. **(c)** GO enrichment analysis of host genes harboring all the splicing outliers, regardless of the association with fusions. The x-axis represents the enrichment significance measured in -(log_10_FDR) and the y-axis shows the GO terms. This figure also refers to Fig. 6.

